# Vaccine Equity in Low and Middle Income Countries: A Systematic Review and Meta-analysis

**DOI:** 10.1101/2022.03.23.22272812

**Authors:** Huda Ali, Anna-Maria Hartner, Susy Echeverria-Londono, Jeremy Roth, Xiang Li, Kaja Abbas, Allison Portnoy, Emilia Vynnycky, Kim Woodruff, Neil M Ferguson, Jaspreet Toor, Katy AM Gaythorpe

**Affiliations:** Imperial College London, Praed Street, London, UK; London School of Hygiene and Tropical Medicine Keppel Street, London, UK; Center for Health Decision Science, Harvard T H Chan School of Public Health Cambridge, USA; Public Health England London, UK

**Keywords:** vaccine, equity, LMIC, systematic review

## Abstract

**Background:** Evidence to date has shown that inequality in health, and vaccine coverage in particular, can have ramifications to wider society. However, whilst individual studies have sought to characterise these heterogeneities in immunisation coverage at national level, few have taken a broad and quantitative view of the contributing factors to heterogeneity in vaccine coverage and impact. This systematic review aims to highlight these geographic, demographic, and sociodemographic characteristics through a qualitative and quantitative approach, vital to prioritise and optimise vaccination policies.

**Methods:** A systematic review of two databases (PubMed and Web of Science) was undertaken using Medical Subject Headings (MeSH) and keywords to identify studies examining factors on vaccine inequality and heterogeneity in vaccine coverage. Inclusion criteria were applied independently by two researchers. Studies including data on key characteristics of interest were further analysed through a meta-analysis to produce a pooled estimate of the risk ratio using a random effects model for that characteristic.

**Results:** One hundred and eight studies were included in this review. We found that inequalities in wealth, education, and geographic access can affect vaccine impact and vaccine dropout. We estimated those living in rural areas were not significantly different in terms of full vaccination status compared to urban areas but noted considerable heterogeneity between countries. We found that females were 3% (95%CI[1%, 5%]) less likely to be fully vaccinated than males. Additionally, we estimated that children whose mothers had no formal education were 28% (95%CI[18%,47%]) less likely to be fully vaccinated than those whose mother had primary level, or above, education. Finally, we found that individuals in the poorest wealth quintile were 27% (95%CI [16%,37%]) less likely to be fully vaccinated than those in the richest.

**Conclusions:** We found a nuanced picture of inequality in vaccine coverage and access with wealth disparity dominating, and likely driving, other disparities. This review highlights the complex landscape of inequity and further need to design vaccination strategies targeting missed subgroups to improve and recover vaccination coverage following the COVID-19 pandemic.

**Registration:** Prospero CRD42021261927

## 1 Background

Vaccination is a vital and effective intervention against disease-related morbidity and mortality, particularly in low- and middle-income countries (LMICs), preventing an estimated 5.1 million deaths from vaccine-preventable diseases annually [1]. The last two decades have seen substantial progress in vaccination coverage alongside a series of global initiatives to decrease vaccine inequity, including the United Nations’s Sustainable Development Goals (SDGs), the creation of Gavi, the Vaccine Alliance, and the development of the Global Vaccine Action Plan 2011-2020 (GVAP). Despite this, progress in global coverage of the three doses of diphtheriatetanus-pertussis vaccination (DTP3), a commonly-used proxy indicator for immunisation performance, has stagnated at 85% since 2010; only 64% of countries achieved the target of 90% coverage or higher [2, 3, 4]. In 2019, an estimated 14 million infants did not receive an initial dose of DTP, highlighting lack of immunisation access and the need to reach individuals and communities missed by routine vaccination activities [3, 4]. Though the Immunisation Agenda 2030 (IA2030) aims to further global immunisation progress and reduce global inequities, the global COVID-19 pandemic has resulted in the further disruption of routine immunisation (RI) and campaign activities, with projections estimating at least 5% fewer fully vaccinated persons (FVPs) and 5.22% more deaths globally, even if IA2030 goals are met [5, 6]. Thus, as the global value of vaccination comes into sharp focus, it highlights a need for a sustained and comprehensive response to maintain equitable, robust, and resilient immunisation services.

Globally, immunisation coverage remains variable both between and within countries, with some populations disproportionately under-immunised. Of the 19.9 million children who had not received the recommended three doses of DTP, 62% resided in just 10 countries^[1]^ [2]. Whilst some heterogeneity and inequality in access is well described at a country level, few examinations have explored the broader mix of factors that contribute to vaccine coverage inequality and access issues. Current studies have highlighted the influence of different demographic, socioeconomic, and access factors contributing to inequalities in vaccine coverage, including maternal and paternal education, wealth, gender, and geography [7, 8]. Notably, however, the directionality of these factors is not always the same, resulting from differences in a country’s approach to immunisation service provision, vaccine introductions, and immunisation maturity, based on the WHO immunisation maturity grid^[2]^ [9, 10].

Populations who are unvaccinated or only partially vaccinated are at higher risk of morbidity and mortality; however, there are further, societal implications of uneven coverage. Subnational coverage disparities resulting from the geographical clustering of disadvantaged subgroups can result in sustained disease transmission and increase the risk of outbreaks [11, 12]. Furthermore, vaccination has been found to increase productivity and cognitive outcomes in children, this in turn can improve social mobility and economic prospects [13, 14]. As such, factors that may contribute to vaccine access inequality may themselves be propagated by low coverage.

Monitoring differences in the inequities of global vaccination coverage is an important step toward tailoring relevant programmes and policies, and allows for direct resource allocation to target missed individuals and communities [12]. We present a systematic literature review on the factors that are associated with vaccine inequality and heterogeneity in vaccine coverage, with a focus on demographic, socioeconomic, and geographic factors, highlighting whether these inequities impact vaccination and subsequent barriers to access. We present and synthesise this work to provide both quantitative and qualitative findings concerning existing immunisation inequities across LMICs. This manuscript uses the term inequality ‘in its neutral sense to denote a measured difference in health between population subgroups, while inequity is used to describe a situation where the distribution of health is unjust, unfair or avoidable’, as defined by the World Health Organization (WHO) [15].

## 2 Methods

Our methods adhere to the guidelines established by Preferred Reporting Items for Systematic Reviews and Meta-Analyses (PRISMA). Our study protocol was registered with PROSPERO (International Prospective Register of Systematic Reviews) under the identifier CRD42021261927.

### 2.1 Searches

The systematic review was conducted using the PubMed and Web of Science databases using the terms (vaccine or vaccination or immunisation or immunization or vaccines) AND (equality or equity or fairness or inequality or disparity) AND (“developing countries” or “low- and middle-income countries” or “middle-income regions” or “low-income regions” or “poorer countries” or LMIC).

### 2.2 Study exclusion and inclusion criteria

Alongside meeting the above search criteria, studies also needed to mention heterogeneity in vaccine access or coverage and be in the English language. No differentiation was made based on whether studies focused on routine immunisation coverage or supplementary immunisation activities. We included studies published between 1974 and the 15th of June, 2021.

Studies with a sole focus on COVID-19 vaccines were excluded, given that we aimed to examine the long-standing inequities in vaccine coverage. We also excluded non-peer-reviewed studies, those that did not include LMICs, duplicates, and studies where vaccination was not mentioned or was not the outcome variable. Similarly, studies with no mention of demographic, geographic, or socio-economic variation in vaccine access or coverage, or no mention of vaccines, were excluded. Finally, we removed editorials, opinion pieces, news articles, or reviews with no empirical data.

### 2.3 Potential effect modifiers and reasons for heterogeneity

We examined variation in vaccine coverage driven by geography, societal structure, political stability, differences in immunisation financing, and the timing of vaccine introduction.

### 2.4 Study quality assessment

Each paper included or excluded was reviewed by two reviewers and reasons for inclusion or exclusion were stated. The Critical Appraisal Skills Programme (CASP) guidelines for qualitative work were used to ensure that each paper chosen, whether qualitative or quantitative, was valid and of high quality [16]. Given the combined analysis, CASP was utilised across studies as an alternative to GRADE, given GRADE does not include metrics for qualitative studies, and in order to maintain consistency. Following data extraction, each study was marked as good, adequate, or poor for each criterion in the CASP guidelines, which was then used to give each study an overall grade. A further quality assessment was performed on quantitative studies included in the meta-analysis to ensure consistency between results and definitions.

### 2.5 Data extraction strategy

Data extraction of quantitative data was performed by one reviewer and verified by another. The aim was to extract the number of fully and incompletely vaccinated individuals in subpopulations disaggregated by geographic, socio-economic, or demographic characteristics. We also collated the countries included in each study, the age range of participants, the vaccines included, the definition of fully vaccinated used in the study, the year of publication, and the year of data collection. Finally, we extracted information on any available contributing factors to examine heterogeneity in access between and within countries. All information was compiled in a spreadsheet accessible by all reviewers.

### 2.6 Data synthesis and presentation

Quantitative data analysis was only conducted on studies that included details on a subpopulation of interest, i.e. the total number of males and females who were fully vaccinated. Prior to analysis, the filtered studies were checked for comparability based on study type and data included. Random effects modelling was performed using the R package *metafor* to produce estimates of the relative risk of full immunisation for each subpopulation; p-values were calculated using a chi-squared test [17]. We used an empirical Bayes estimator for the level of heterogeneity and weighted by the size of the population. We performed five hypothesis tests ^[3]^, each evaluating the null hypothesis that the risk of full immunisation is the same across two levels of a socio-demographic characteristic of interest. We declared significance if a p-value was below 0.01, which was chosen after applying the Bonferroni correction to our starting significance level of 0.05 and taking into the account the five tests. Additional information is provided in the supplementary index. Adjusted risk ratios were not calculated as it was not possible to link covariates for the majority of the available data. Analyses were conducted using R version 4.0.3. Data and code are available from https://github.com/mrc-ide/vaccinequity_litreview.

## 3 Results

### 3.1 Overview

A total of 1573 potential studies were identified through a literature search, and 210 duplicates were removed before screening. Titles and abstracts for 1363 studies were screened. Of these, 286 met the inclusion criteria for full-text evaluation. Finally, 108 studies remained after excluding 178 studies due to the following criteria: (1) Vaccination was not an outcome variable; (2) There was no mention of heterogeneity or vaccine inequality; (3) The study was an editorial/comment/opinion or news article with no empirical data. The PRISMA flow diagram is shown in figure 1.

**Figure 1:**
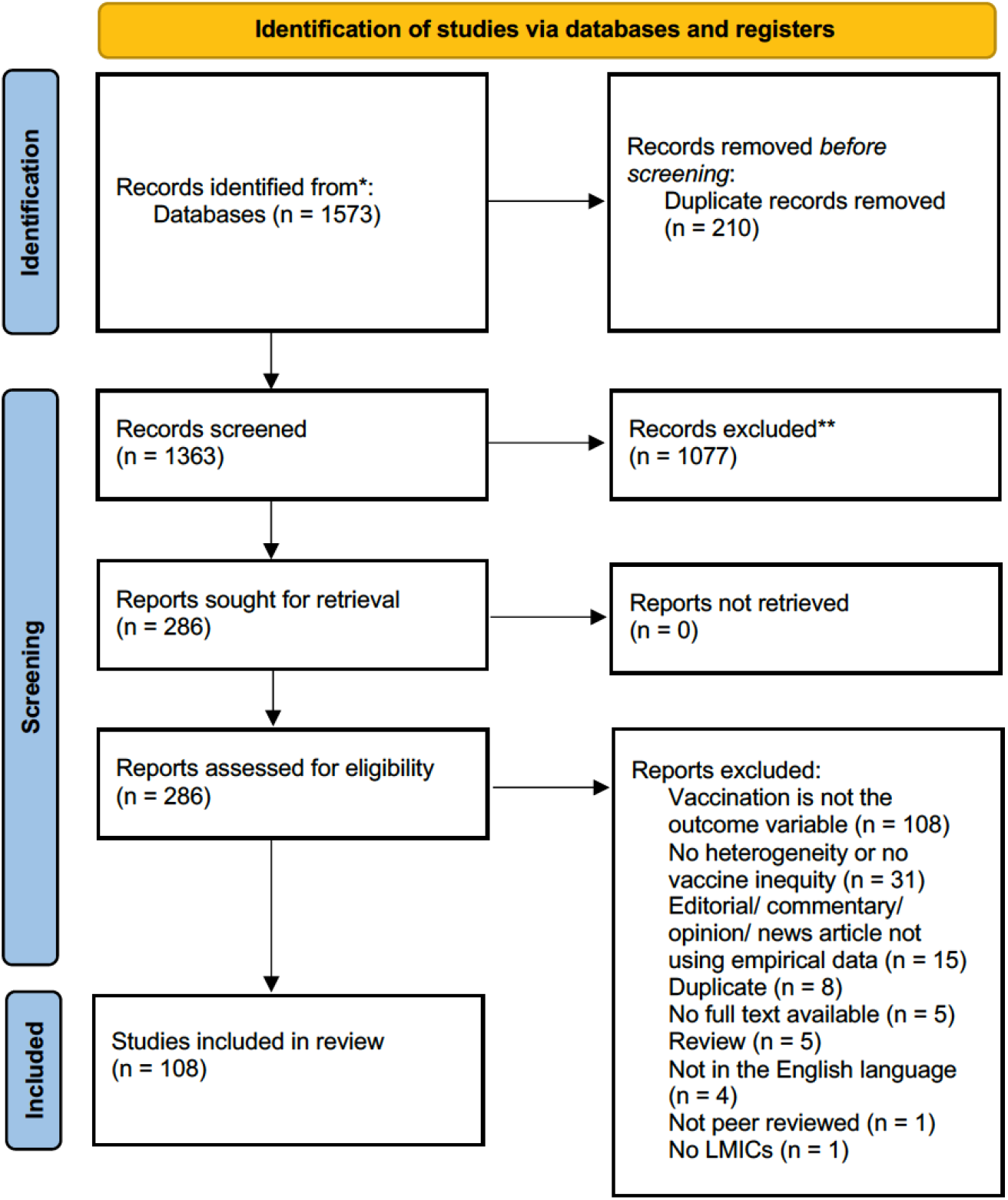
PRISMA flow diagram showing number of studies included at each review stage [18].

#### 3.1.1 Study characteristics

Where studies examined only one country, India (24 studies) was most frequently considered, followed by China (9) and Bangladesh (8). While generally more individual studies focused on the Asian continent, the African continent was highly represented in studies covering multiple countries. All studies, covering 132 countries in total, are included in figure 2a including those covering multiple countries [19, 8, 20, 21, 22, 23, 3, 24, 25, 26, 27, 7, 11, 28, 12, 29, 30, 31, 32, 33, 34].

**Figure 2:**
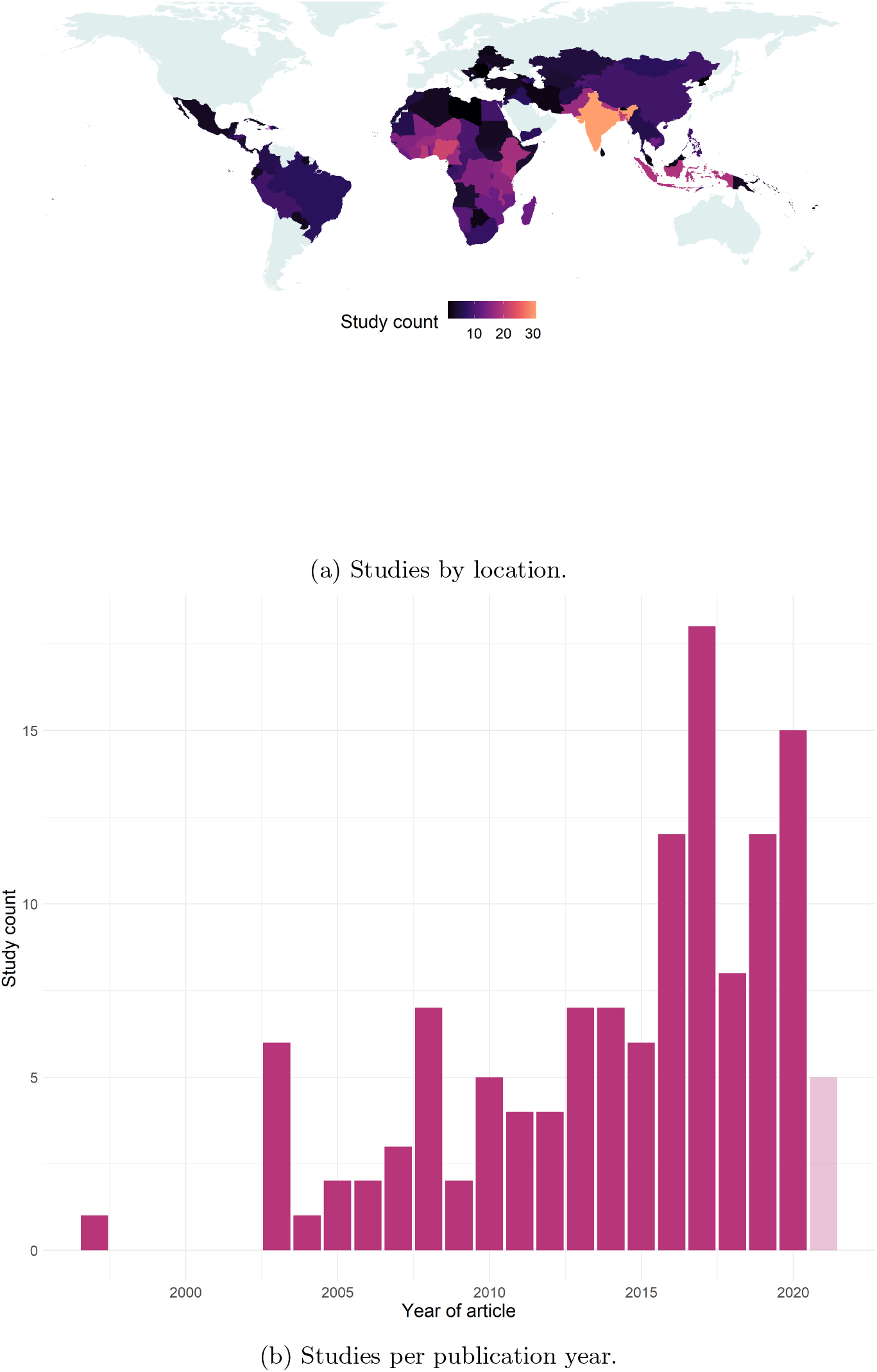
Geographic and temporal distribution of studies. 2021 is shown with transparency as the search was conducted mid-way through the year and thus the number of potential studies is likely to be incomplete.

The studies were not evenly distributed over time, with an increasing number of included articles published in recent years, see figure 2b. The earliest included study by publication date was released in 1997 [35], though the earliest year of data collected was from 1986 [22]. The most recent data was from 2019 [3, 11]. Most studies examined vaccination in children under 5 years old, only 2 included vaccination coverage in adults [36, 37] and three omitted a clear description of age, see figure A.1 for all age ranges and details. A summary table of all included definitions of fully vaccinated can be found in the supplementary material, table A.3.

#### 3.1.2 Study quality

The included studies varied against the CASP guidelines; however, the majority achieved the highest grade (good), with only three achieving the lowest (poor). These three lacked some details on methodology and ethical considerations, but contained clear statements on their findings. All studies were included in the thematic analysis, or, where they contained relevant quantitative information, the meta-analysis. All study grades are included in the supplementary information and code.

For studies including quantitative information, we further reviewed their definition of vaccinated to ensure consistency; this resulted in the removal of one study, Uthman et al., as it was not feasible to compare fully vaccinated individuals against incompletely vaccinated individuals [38]. This removal did not change the significance of the relative risk estimates, and results of the meta-analysis with this study included are shown in supplementary table A.2.

### 3.2 The effect of inequality in vaccination coverage on the impact of vaccination

We found relatively few studies that examined how inequality in vaccination coverage among population subgroups affected the overall benefits of vaccination in that population. This may be due to the framing of our search queries i.e. excluding studies where vaccination was not the outcome variable. However, some points held across the countries studied, particularly concerning the prevalence of full vs partial immunisation and targeting interventions. Inequities such as wealth, education, and geographic access to health services have been linked to increased risk of dropout in vaccine courses and higher risk of infection from pathogens such as Hepatitis B [37, 39, 40, 41, 42]. Similarly, less advantaged populations may see a delay in full immunisation leading to increased potential risk of infection [43]. This disruption of full immunisation can lead to the vaccination itself appearing less effective as individuals are missed or not effectively immunised. Conversely, vaccination occurring in areas of low or zero coverage can appear more impactful as the baseline level of protection is lower; studies including Helleringer et al. and Portnoy et al. note this especially in disadvantaged populations who have been targeted for measles supplementary immunisation activities (SIAs) [27, 33, 29].

### 3.3 Geographic variation in vaccination coverage

Information on geographic heterogeneity in vaccination coverage mainly focused on discrepancies between urban and rural areas and the reasons behind these differences. The overview is a mixed picture — in some countries, such as China, coverage and probability of full immunisation are higher in rural areas. However, in Ethiopia, the opposite relationship is seen. This variation is highlighted in figure 3.

**Figure 3:**
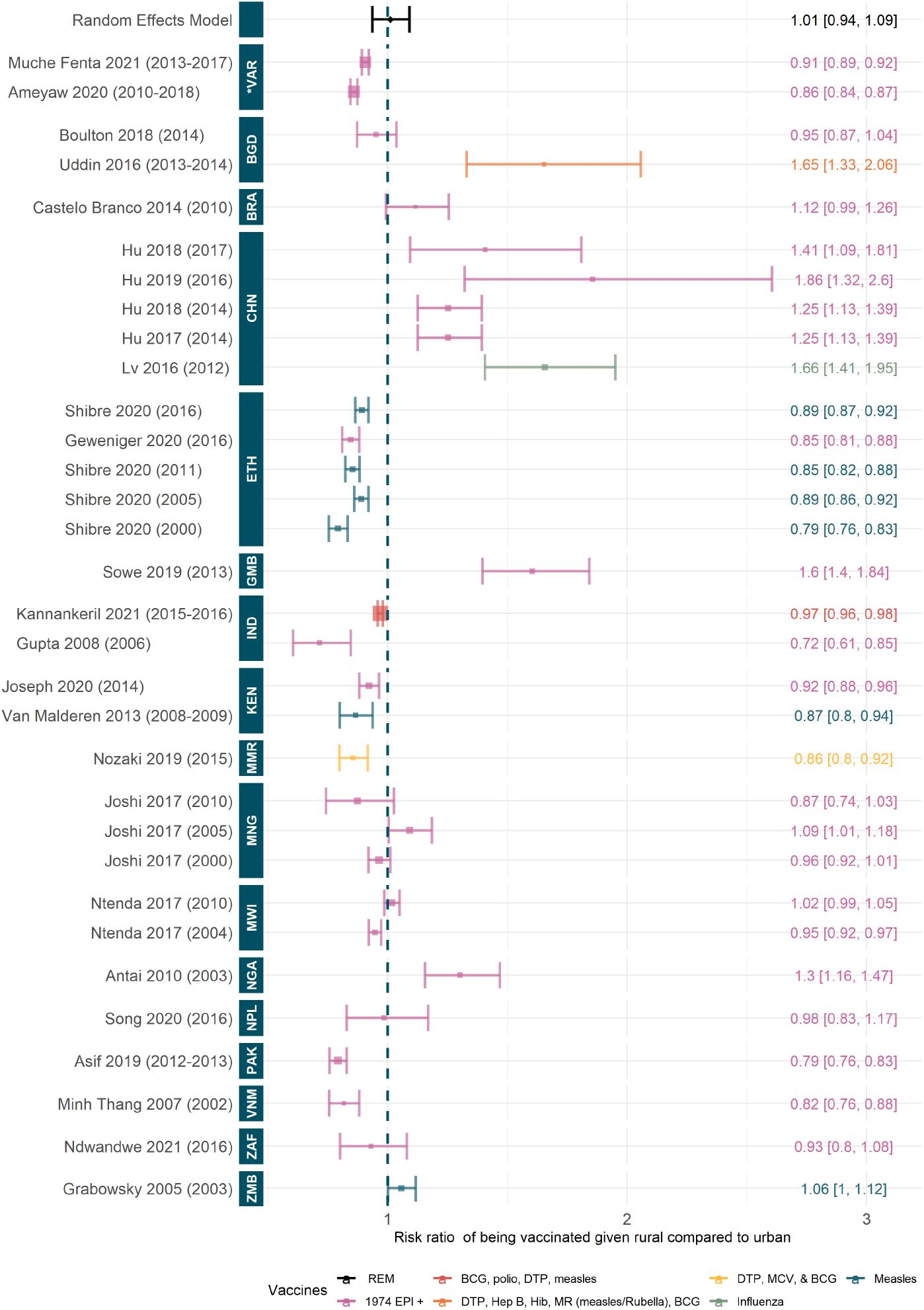
Relative risk to being fully vaccinated given rural compared to urban dwelling. Random effect model estimate is shown in black and p-value of the fit is not significant (0.78). Colours denote type of vaccines considered, see table A.3 for full details. Studies are ordered by the year of data, shown in brackets, and country of data. ISO codes are: ZMB=Zambia, ZAF=South Africa, VNM=Vietnam, PAK=Pakistan, NPL=Nepal, NGA=Nigeria, MWI=Malawi, MNG=Mongolia, MMR=Myanmar, KEN=Kenya, IND=India, GMB=Gambia, ETH=Ethiopia, CHN=China, BRA=Brazil, BGD=Bangladesh and *VAR=Various. Studies included: [92, 93, 24, 19, 88, 82, 80, 86, 94, 95, 69, 75, 76, 62, 96, 50, 55, 49, 97, 46, 48, 53, 70, 73].

Vaccination coverage was higher in rural areas than urban areas in China, the Gambia, Mauritania, Nigeria, Swaziland, and Uzbekistan. In China, overall coverage was generally high and most provinces have reached Gavi targets [44]; Studies by Cui et al., Hu et al., Xie et al., and Lv et al. have suggested that rural residents have better access and relationships with their healthcare providers, leading to higher vaccination coverage [45, 46, 47, 48, 49]. In the Gambia, coverage is higher in rural areas but varies substantially in completion of vaccination [50, 51]. Finally, Restrepo-Mendez et al. noted that three countries — Mauritania, Swaziland and Uzbekistan — had significant pro-rural vaccination coverage rates(i.e. full immunisation was more prevalent among children living in rural areas), but this effect was in the minority of countries examined. [7].

Coverage, or the prevalence of full immunisation, was higher in urban areas of Brazil, Cameroon, Ethiopia, India, Madagascar, Malawi, Myanmar, Tanzania, Pakistan, and Vietnam. Brazil achieved high vaccination coverage with no large differences between regions; despite this, living in rural areas was found to be associated with incomplete vaccination [52, 53]. In Ethiopia, there were significant urban-rural differences in coverage, although coverage levels in rural areas were increasing more quickly than urban; these differences contribute to Ethiopia having one of the lowest overall vaccination coverage rates in sub-Saharan Africa [54, 55, 56]. India has seen huge improvements in reducing the number of zero-dose children and heterogeneity in coverage; however, there are still significant urban-rural differences in coverage and the change in number of fully immunised children has stagnated [57, 58, 59, 40, 60, 61, 62, 63]. Other factors may be influenced by, and influence, the urban-rural differences. Sissoko and Prusty especially noted that rural living may have a protective effect for vaccination when controlling for other factors, and that the pro-male gender divide in coverage in rural areas is greater than that in urban areas of India [64, 65]. In Madagascar and Malawi, coverage was higher near the capital and full vaccination was associated with urban areas; yet both urban and rural areas were heavily affected by access to health services [66, 67, 68, 69]. In studies that examined multiple countries, the consensus was that rural areas generally have lower overall vaccination coverage and higher prevalence of incomplete vaccination than urban areas [19, 3, 24, 7, 28, 31].

While vaccination coverage is generally higher in urban areas, this can mask further heterogeneity. Low coverage in urban informal settlements and among the urban poor was highlighted in Bangladesh, Burkina Faso, Kenya, and Ghana. In Bangladesh, urban children were more likely to be fully immunised than rural; however, coverage in urban informal settlements was lower than the urban average, leading to a statistically insignificant difference between urban and rural coverage overall [70, 71, 72, 73]. In Burkina Faso, Kenya, and Ghana, the protective effect of urban living led to a higher probability of full immunisation coverage but pockets of urban poverty saw low coverage and diluted the urban-rural difference [42, 74, 75, 35, 43]; similarly, targeted vaccination in Kenya led to protected subpopulations in rural areas, balancing the urban-rural difference for measles [76].

Negligible or contrasting differences between urban and rural settings were noted in Cambodia, Indonesia, and South Africa with general geographic heterogeneity suggested in Afghanistan, Mozambique, Nepal, and Togo [77, 41, 78, 79, 80, 81, 38, 82, 83, 84, 85, 86, 87, 88, 89]. Dropout of vaccination driven by transport costs and access was highlighted in South Africa and Uganda, despite mitigation through outreach activities [90, 91].

#### 3.3.1 Quantitative synthesis of rural-urban differences in vaccination coverage

No significant pooled effect was found for the likelihood of being fully vaccinated given residing in rural compared to urban areas (Figure 3). This is due to the substantial variation between countries. In China, the Gambia, Nigeria, and Zambia, for all years of data and included studies, there is a significant relative benefit to being vaccinated given living in a rural area compared to urban settings [49, 97, 46, 48, 50, 80, 92], whereas in Ethiopia, India, Kenya, Myanmar, Pakistan, and Vietnam, the opposite relationship is seen [55, 56, 62, 96, 76, 75, 94, 82, 93]. Similarly, Ameyaw et al. and Muche Fenta et al., who examined multiple countries in Africa, found significant relative risks to being vaccinated given residing in rural settings compared to urban areas [19, 24]. Other included studies found no significant effect or found contrasting effects to other studies in the same country.

### 3.4 Demographic variation in vaccination coverage

We explored demographic heterogeneity in vaccination coverage through a variety of factors, including gender, age, birth order, religion, and ethnicity, and/or caste. Our overall findings were mixed, with these factors varying significantly by country, region, and year.

Immunisation coverage differences by gender varied broadly globally, but pro-male gender disparities were identified in Bangladesh, Brazil, Cameroon, India, and Nepal. In Bangladesh and Nepal these differences were minimal, though this disparity was found to increase with poverty or lower maternal education in Bangladesh [72, 87]. In Brazil, the overall compliance with the recommended hepatitis B vaccination schedule was found to be associated with gender [37], while in Cameroon, gender disparities were found to favour males, though this trend reversed with time [98]. The greatest differences in immunisation coverage as a result of gender were seen in India — here, females were more likely to be completely unvaccinated or have incomplete vaccination statuses. [57, 58, 62, 25]. Interestingly, this trend did not hold when considering complete immunisation system failure, defined by Gaudin as when “the infrastructure to provide immunization [was] not in place, affecting all groups” [59], and in some instances, varied when other demographic factors, including religion and caste, were taken into account [64]. The reverse trend, in which full immunisation coverage was greater in girls than boys, was seen in just two studies in which either the overall coverage disparities were low or in which other inequities were taken into account, suggesting gender disparities in India may be influenced by other demographic and socioeconomic outcomes [99, 64].

No significant immunisation coverage disparities by gender were observed in Cambodia, Ethiopia, Indonesia, Madagascar, Malawi, Mongolia, Mozambique, Myanmar, or Pakistan [41, 55, 56, 78, 66, 95, 68, 84, 83]. In Madagascar, though no overall gender differences were observed, results did suggest that the influence of paternal educational attainment on immunisation coverage was greater among males [66]. In Myanmar, though gender was not found to be significant, slightly higher coverage rates were observed in boys compared to girls [84].

Insights on cultural or policy-level differences contributing to this relationship were provided by studies in Afghanistan and China. In Afghanistan, though the direct relationship of gender on immunisation coverage was not provided, the overall lack of female autonomy and its limitations on healthcare access were described, with implications towards the accessibility of immunisation services for women and girls [77]. In China, while one study found females to be less likely to be vaccinated, a study on free influenza vaccination in the elderly found no correlation with gender, suggesting the disparity may be age dependent [36, 48].

In studies that examined multiple countries, the picture was mixed, with some finding no significance between genders [19, 3] and others finding vaccine- or country-specific differences that favoured males over females [8, 21, 7]. Of the papers that found differences favoring males over females, Arsenault et al. found only six countries, Lesotho, India, Burkina Faso, Gambia, Côte d’Ivoire, and Pakistan, had statistically significant differences in DTP3 coverage between genders [8]; Bonu et al. found gender differentials for Bangladesh, Côte d’Ivoire, India, Malawi, Nepal, and Rwanda [21]. Similarly, Restrepo-Méndez et al. found small gender-related differences, with the gender disparity reaching statistical significance in Azerbaijan, Belize, India, Mali, and Somalia [7].

Ethnicity or caste were found to be significant factors contributing to immunisation coverage in Bangladesh, Cambodia, China, Ethiopia, Gambia, India, Namibia, Nepal, Pakistan, and Vietnam with lower immunisation rates among ethnic minorities [71, 41, 97, 56, 51, 100, 93, 82, 99, 86, 85]. In India and Nepal, caste was found to be a highly significant contributor to immunisation coverage, with children from lower castes less likely to be immunised [99, 100, 86, 85]. Though the gap for oral polio vaccine (OPV) coverage in India declined as a result of caste over time, it remained significant at the bivariate level, when results were unadjusted for other potential confounding factors [100]. Notably, the degree of immunisation inequality was found to be less associated with ethnicity in just one country, Kenya — and was suggested to be the result of confounding inequalities as a result of wealth and parental education [74].

Religion remained an important demographic factor in Burkina Faso, Cameroon, Ethiopia, Kenya, India, and Nigeria [42, 98, 56, 101, 100, 57], though in Nigeria this factor was only identified when results were stratified by wealth [101]. In India, children who were part of the Muslim minority were less likely to be vaccinated, and notably this demographic factor further increased the gender disparity in immunisation coverage; i.e. Muslim females were significantly more disadvantaged than their Hindu male counterparts [100, 57].

Birth order or greater family size contributed to changes in immunisation coverage in China, India, Indonesia, Kenya, Mozambique, Nigeria, Pakistan, Swaziland, and Tanzania, though the direction of impact varied. In India, Indonesia, Kenya, Mozambique, Nigeria, and Pakistan, children of a higher birth order or within families of three of more children had a greater risk of incomplete vaccination [102, 79, 75, 84, 80, 83]. For China, Swaziland, and Tanzania, the trend was reversed, with children from larger families having higher full vaccination coverage [97, 46, 103, 104]. Some of these trends are likely to be linked to the age of the mother; in Cameroon, China, Kenya, and Nigeria the increasing age of the mother was tied to increased vaccination coverage [98, 46, 75, 80]. Myanmar was the only country to find no association between the number of children in the household and vaccination coverage [94]. Among studies that examined multiple countries, immunisation coverage was higher among children of lower birth orders, and in a study examining sub-Saharan Africa, lower among children with older mothers compared to younger mothers [19, 24]. Finally, only one study examining the effect of marital status on immunisation found an association [24].

In summary, demographic heterogeneity in vaccination coverage as a result of gender, age, birth order, religion, and ethnicity and/or caste generally found broad variances attributed to country or region-specific cultural or policy differences, though data for several countries was often limited and suggested other confounding factors.

#### 3.4.1 Quantitative synthesis of gender differences in vaccination coverage

We found a significant relative risk to being fully vaccinated given female compared to male of 0.97 (95%CI [0.95, 0.99]) suggesting females are 3% (95%CI [1%,5%]) less likely to be vaccinated than their male counterparts, see figure 4. However, there is some heterogeneity when we examine individual countries and studies. In the majority of included studies, the relative risks are not significant and the confidence intervals span one; a small number of studies or datasets suggest a significant relative risk [95, 57, 50, 48, 53, 72, 71] whereas there are no studies that suggest a significant relative benefit to being vaccinated given female. We also note some changes over time for Bangladesh, China and Ethiopia where studies based on later data have a relative risk closer to one than earlier studies. In Ethiopia, the latest data suggests a central relative risk over one although in all cases, the ranges span one.

**Figure 4:**
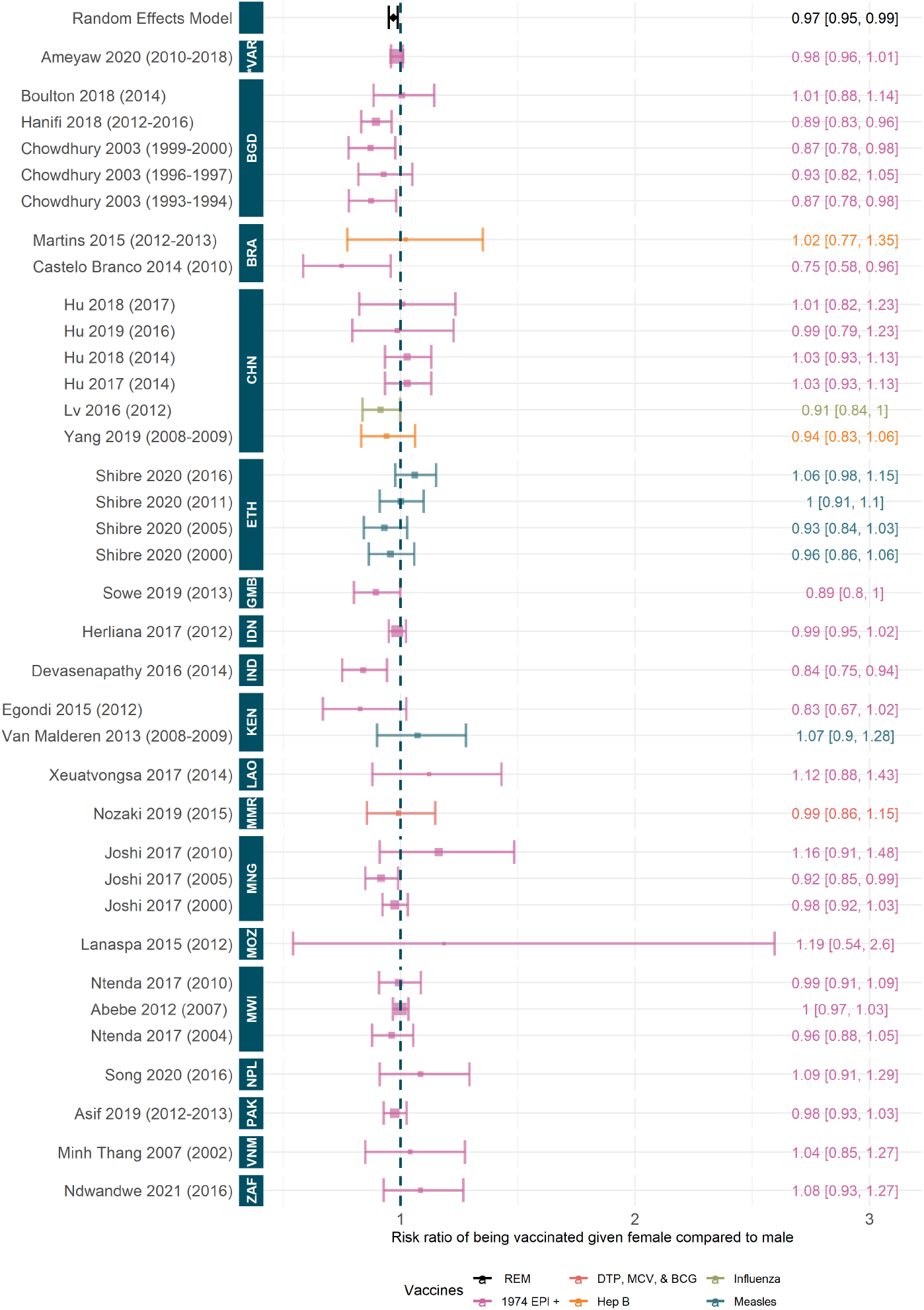
Relative risk to being fully vaccinated given female compared to male. Random effect model estimate is shown in black and p-value of the fit is significant (1.0×10^−3^). Colours denote type of vaccines considered, see table A.3 for full details. Studies are ordered by the year of data, shown in brackets, and country of data. ISO codes are: ZAF=South Africa, VNM=Vietnam, PAK=Pakistan, NPL=Nepal, MWI=Malawi, MOZ=Mozambique, MNG=Mongolia, MMR=Myanmar, LAO=Laos, KEN=Kenya, IND=India, IDN=Indonesia, GMB=Gambia, ETH=Ethiopia, CHN=China, BRA=Brazil, BGD=Bangladesh and *VAR=Various. Studies included: [97, 86, 55, 88, 46, 94, 105, 57, 70, 50, 37, 48, 84, 79, 72, 74, 82, 69, 95, 53, 19, 106, 76, 68, 93, 71].

### 3.5 Socioeconomic variation in vaccination coverage

Information on socioeconomic heterogeneity in vaccination coverage focused predominantly on household wealth, maternal and paternal education, and occupation. These findings describe increasing coverage among families in higher wealth quintiles and in families with increased levels of maternal or paternal education, though a handful of countries described a mixed picture. Outcomes that favored pro-poor inequalities and low educational attainment were likely the result of confounding by urban and rural differences between country settings; negligible outcomes may be the result of differences in the measures of economic status, or, in some settings, well-established public immunisation programmes. Socioeconomic factors often retained their significance in adjusted analyses, suggesting it is a significant driver of immunisation differences in LMICs.

Immunisation coverage was lower among the poorest wealth quintiles in Bangladesh, Brazil, Burkina Faso, Cambodia, Gambia, Ghana, India, Indonesia, Kenya, the Kyrgyz Republic, Madagascar, Malawi, Mongolia, Mozambique, Myanmar, Nigeria, South Africa, and Swaziland [42, 51, 66, 67, 68, 69, 94, 103, 101, 80, 81, 107, 38]. With DTP3 dropout as a measure of incomplete vaccination status, Cambodia saw a decrease in pro-rich inequality over time, but coverage differences between wealth indices were still significant [39, 41]. In Ghana, socioeconomic predictors were thought to account for regional variation in coverage rates; wealth quintile was additionally found to be associated with a delay in vaccination among children [35, 43]. This was similarly observed in India, where increasing wealth was found to be a significant predictor of full immunisation coverage and lower zero-dose prevalence, even before and after campaign implementation, and was also found to influence timely, age-appropriate immunisations [108, 99, 40, 100, 102, 57, 109, 110, 62, 60, 64]. Increasing gaps in immunisation coverage related to socioeconomic status in India may indicate a further widening in the rich-poor gap in child care services [111]; however, in Bangladesh these gaps were narrowing by relatively faster improvement in poorer wealth quintiles [112]. While Sissiko et al. found poorer household wealth was a stronger predictor of completely unvaccinated in rural settings but not urban, Prakash et al. continued to find inequities among the urban poor when compared to the non-poor, especially for DTP and measles [65, 113]. Differences in immunisation inequalities associated with wealth in urban versus rural settings were also observed in South Africa, where the difference has been suggested to be the result of healthcare access; urban township sites have been thought to increase the use of public health services among the urban poor when compared to the rural poor [88, 90]. In Indonesia and Kenya, the socioeconomic inequality in immunisation was found to be especially correlated to measles vaccination uptake alongside the association with increased odds of full vaccination, though Kenya observed changes to this correlation over time [78, 79, 74, 76, 75]. In Mongolia, economic status was found to only be significant in pockets of low overall coverage, ultimately losing significance as coverage improved [95].

Outside predictors of socioeconomic status were found to have an impact in observed pro-rich inequalities in some countries. In Bangladesh, full immunisation coverage was higher for households above the poverty line; a proxy indicator, “self-rated food security status,” was similarly associated with higher immunisation coverage in that chronically “food deficit” households had a nearly 50% lower coverage rate than those in “surplus” households [70, 71, 72]. In Brazil and South Africa, inadequate housing was found to be a strong predictor of incomplete vaccination associated with increasing income disparity, and wealth inequality was found to further impact the outcome of hepatitis B vaccination in Brazil [37, 53, 114], while in Mozambique and Malawi, families with safe water had an increased likelihood of vaccination coverage [84, 68].

In studies that examined multiple countries, children from families in the richest wealth quintiles had a greater likelihood of full immunisation coverage, with notable inequalities in DTP3, OPV, and MCV1 coverage and in MCV, DTP1, and DTP3 dropout rates [19, 8, 20, 21, 22, 3, 25, 26, 27, 7, 28, 12, 30, 31, 24, 115]. Children from poorer households were additionally more likely to have an increased zero-dose prevalence, and in some countries, lower participation rates in SIA [27, 3], though this varied [33].

The magnitude of pro-rich inequality in immunisation coverage has been shown to vary by the specific measure of economic status used [32]. Studies in China, Ethiopia, Namibia, Nepal, Pakistan, Tanzania, Thailand, and Vietnam described a mixed picture of the association between wealth and immunisation coverage. In China, while some studies described lower coverage among lower wealth quintiles, changes in immunisation uptake between quintiles were small [45, 47], and in Yang et al., unobserved [106]. In the case of influenza vaccination among the elderly, the reverse was observed, in which coverage showed a pro-poor distribution [36]. In Ethiopia, two studies described wealth-based inequities displaying a pro-rich distribution [55, 56], with one describing a decrease in this inequality over time [54]. Wuneh et al., however, found no differences by wealth in rural Ethiopia [116]. In Nepal, Pakistan, and Tanzania, associations in the disparity of vaccination coverage among wealth quintiles was found to change over time, with Nepal and Pakistan displaying a decreasing disparity and Tanzania showing increases [85, 86, 87, 82, 117, 118, 83, 104]. In Vietnam and Thailand, studies found conflicting associations with immunisation and wealth [93, 119, 25]. One study on Namibia described a pro-poor association in vaccination coverage, though this was described as confounded by the urban-rural divide, suggesting that no association would exist without regional differences [51].

Several studies additionally examined the association between parental occupation (often used as a proxy for household wealth) and children’s immunisation status. In Nigeria and Pakistan, the children of nonworking mothers had higher rates of immunisation, while in Bangladesh, paternal occupation had a higher, positive, association with childhood immunisation [71, 80, 82]. In Laos, neither maternal nor paternal occupation showed any association with vaccination status, potentially due to the free expanded programme on immunization (EPI) [105]. Similarly, we found no significant association between maternal marital status and full immunisation status, see figure A.2 in the supplementary material.

Parental education further contributed to socioeconomic heterogeneity in immunisation coverage globally, with increasing parental education contributing to improved coverage outcomes in Burkina Faso, Cambodia, China, Ethiopia, Gambia, Ghana, India, Indonesia, Kenya, Laos, Madagascar, Malawi, Namibia, Nepal, Nigeria, Pakistan, Tanzania, and Togo. While general trends for increasing vaccination coverage in children was observed with increased parental education attainment in Burkina Faso, Gambia, Kenya, Laos, Madagascar, Namibia, Pakistan, Tanzania, and Togo, [42, 28, 74, 75, 76, 105, 66, 82, 104, 89], some countries observed additional differences in coverage by vaccine. In Cambodia, differences in education level resulted in differences in DTP3 coverage, while in Ethiopia and Indonesia, uptake in measles immunisation varied by educational status [41, 55, 56, 54, 78, 79]. Indonesia additionally found differences in BCG, OPV3, and DPT3 vaccination coverage [120]. Among the elderly in China, educational status was found to be a stronger predictor of influenza vaccine uptake than economic status [36], though education was found influential for all vaccines [97, 47]. Ghana additionally found an association between increased parental education and adherence to pentavalent diphtheria-pertussistetanus-hepatitis B-haemophilus influenzae B (DTP-HepB-Hib) and polio vaccine schedules [43, 35]. In India, increasing maternal and paternal education was found to significantly improve immunisation coverage of children [108, 99, 40, 65, 62, 109, 60]. The odds of vaccination were lower in children born to illiterate mothers, in which illiteracy is used as a proxy measure to educational attainment [57, 102, 110]. Similar associations between education or literary and vaccination coverage were found in Malawi and Nigeria [69, 101, 81, 38, 68]. While Nepal found associations between parental education and full immunisation coverage, the association compared to other studies was notably smaller [87, 85]. Studies examining multiple countries found that parental educational attainment, especially formal education, and literacy contributed to a greater likelihood of being fully immunised, especially for DTP3 [8, 21, 22, 24].

Negligible or negative associations between educational attainment and vaccination coverage in children were only observed in Cameroon, Swaziland, and Thailand and a mixed picture was seen in Bangladesh, Brazil, and Mongolia. While the reasons for this were not given for Cameroon or Swaziland, in Thailand, this difference has been suggested to be the result of better service coverage in rural areas, primarily by district health systems, than in urban areas [98, 103, 119]. In Bangladesh, increasing maternal education was found to improve coverage rates, especially for MCV1 by Gao et al. and Chowdhury et al., but Boulton et al. found education to be a non-significant factor in determining vaccination outcomes [25, 71, 70]. In Brazil, studies found that education increased vaccine compliance, but also had no overall effect [53, 52], while in Mongolia, the effect of education was only influential in areas of low overall coverage [95].

#### 3.5.1 Quantitative synthesis of maternal education differences in vaccination coverage

We estimate the relative risk to a child being vaccinated given their mother has no formal education compared to having any education level ie. primary or above, as 0.73 (95%CI [0.64, 0.84]); this model fit is significant, see figure5. This result implies that children are 27% (95%CI [16%,36%]) less likely to be fully vaccinated if their mother has no formal education. The results are consistent across the included studies on a country-level; only studies in Mozambique, Mongolia, and Kenya contain confidence intervals that span one, suggesting no significant differences by education status. Only two studies, in Malawi and the Gambia, implied a benefit of lack of maternal education on vaccination status [84, 95, 68, 74, 76, 50]. All other studies of datasets (n = 14) suggest a strongly negative influence of lack of maternal education on child immunisation [93, 24, 19, 82, 101, 86, 69, 75, 55, 70].

**Figure 5:**
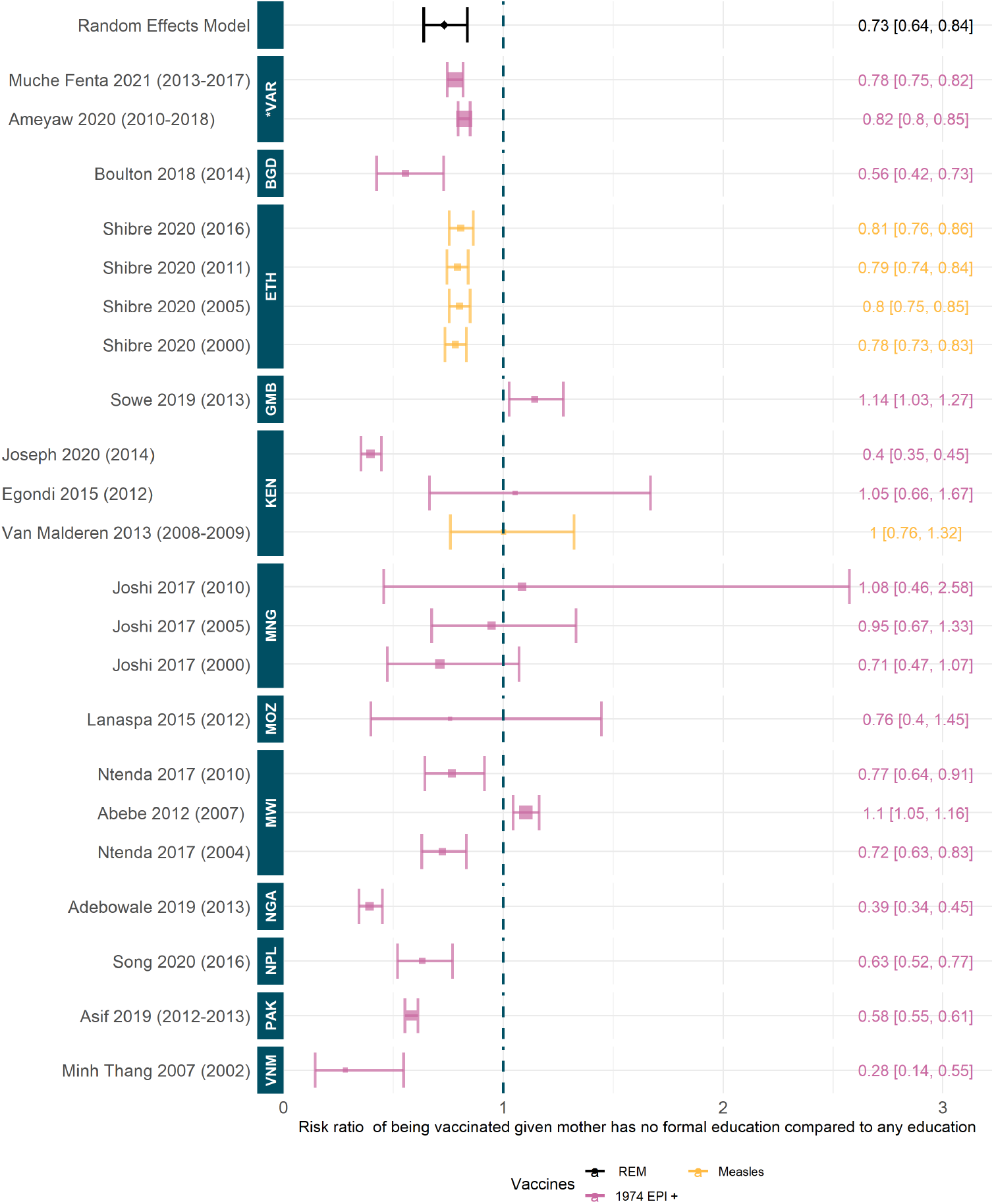
Relative risk to being vaccinated given mother has no formal education compared to mother having primary education or higher. Random effect model estimate is shown in black and p-value of the fit is significant (6.62×10^−6^). Colours denote type of vaccines considered, see table A.3 for full details. Studies are ordered by the year of data, shown in brackets, and country of data. ISO codes are: VNM=Vietnam, PAK=Pakistan, NPL=Nepal, NGA=Nigeria, MWI=Malawi, MOZ=Mozambique, MNG=Mongolia, KEN=Kenya, GMB=Gambia, ETH=Ethiopia, BGD=Bangladesh and *VAR=Various. Studies included: [86, 55, 75, 70, 50, 24, 101, 84, 74, 82, 69, 95, 19, 76, 68, 93].

#### 3.5.2 Quantitative synthesis of wealth quintile differences in vaccination coverage

We estimated a relative risk of 0.73 (95%CI [0.63, 0.84]) to being vaccinated if in the poorest wealth quintile compared to richest; this fit is significant. Figure 6 shows the log relative risks. This suggests that individuals in the poorest subpopulations are 27% (95%CI [16%,37%]) less likely to be fully vaccinated than those in the richest. This result is consistent across the vast majority of included studies and datasets, only Nepal, the Gambia, China, and Brazil have studies suggesting a negative influence or no significant trend [86, 50, 46, 48, 52]. We do note some variation over time in Ethiopia and China. Later studies in Ethiopia suggest a risk ratio closer to one (central estimate 0.63 compared to 0.4) whereas later studies in China suggest widening pro-rich inequality (central estimate 0.5 compared to 1.44).

**Figure 6:**
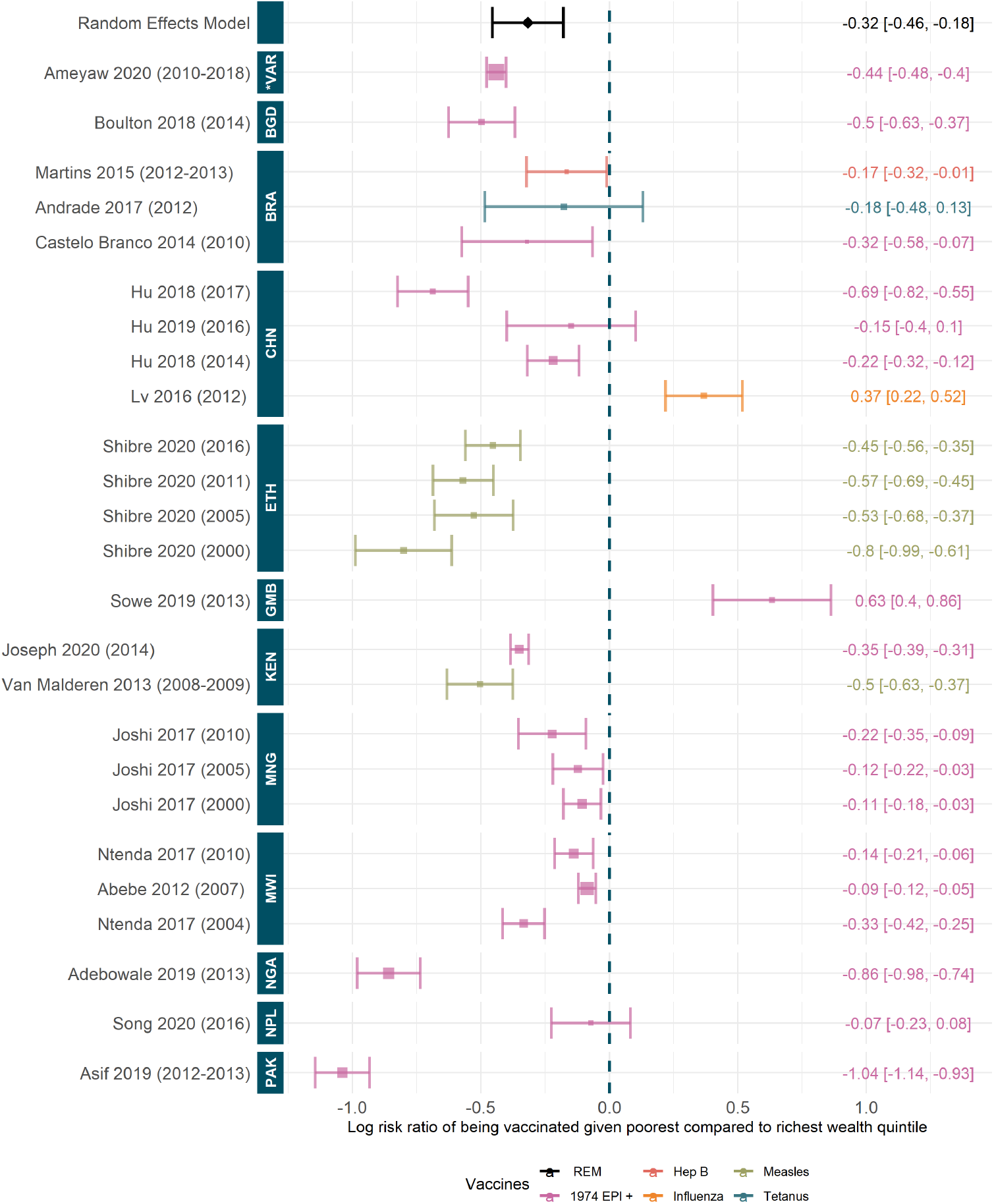
Log relative risk to being vaccinated given poorest compared to richest wealth quintile. Random effect model estimate is shown in black and p-value of the fit is significant (7.51×10^−6^). Colours denote type of vaccines considered, see table A.3 for full details. Studies are ordered by the year of data, shown in brackets, and country of data. ISO codes are: PAK=Pakistan, NPL=Nepal, NGA=Nigeria, MWI=Malawi, MNG=Mongolia, KEN=Kenya, GMB=Gambia, ETH=Ethiopia, CHN=China, BRA=Brazil, BGD=Bangladesh and *VAR=Various. Studies included: [97, 86, 55, 46, 75, 70, 50, 101, 37, 48, 82, 52, 69, 95, 53, 19, 76, 68].

### 3.6 Barriers to vaccine access in Low and Middle income countries

Access to vaccination can be limited by factors such as travel time and cost, safety concerns where there is political instability, the challenges of balancing seeking healthcare and work, and finally, vaccine hesitancy and awareness of immunisation services. In Ethiopia, India, Kenya, Madagascar, Vietnam, and Zimbabwe, travel time to a vaccination facility was strongly related with coverage achieved; for example, in Kenya, 78% of those who lived less than one hour from a healthcare facility were fully vaccinated, compared to only 60% among those who did not. [55, 40, 75, 121, 93, 19]. In Afghanistan and across 45 Gavi-supported countries, vaccination coverage did not improve in areas of political instability even with increased vaccinators and health facilities [77, 8]. Awareness on the benefits of vaccination and autonomy in health decisions can improve coverage — for example, in Bangladesh more autonomy for women led to an improvement in coverage from 78.8% to 86.1% [70]. However, where access is infeasible, or there are competing demands such as work, awareness is insufficient on its own [97, 50]. Migratory and nomadic communities may be further affected by travel time to vaccination centres and outreach although the individual characteristics of these and informal settlement populations can vary substantially [42, 56]. Similarly, the same distance to a health facility may translate differently between wealth quintiles if travel cost and time is a barrier even when immunisation itself is free of charge [122].

## 4 Discussion

We identified 108 studies spanning between 1997 to 2022 that provided information on the factors contributing to vaccine inequality in LMICs, with the most individually studied countries being India, China, and Nigeria. Whilst relatively few of the included studies examined the implications of inequality in vaccination coverage among different population subgroups on the overall impact of vaccination in that population, we found that improvements in wealth, education, and geographic access were linked with reduced dropout from vaccination programmes and reduced delays in reaching full immunisation. This leads to decreased risks of morbidity and mortality. However, disadvantaged populations have also been noted to contain more zero-dose children, meaning disadvantaged populations have greater disease burdens and mortality; as such, targeted interventions such as SIAs have a larger perceived impact than they would on a partially immunised population [33]. This study is the first review to assess vaccine inequality amongst a wide range of LMICs, with this literature review painting a much broader and nuanced picture of vaccine inequality amongst different countries, a key strength.

Geographic variation was noted in almost all regions with particular emphasis on the urban-rural divide. We found a mixed picture of how this divide translated into vaccination coverage. While the majority of countries saw a higher coverage achieved in urban settings, particularly regarding complete immunisation, this is likely driven by access to available clinics both in travel time and cost. Conversely, the urban poor or those living in informal settlements were less likely to reach full immunisation coverage than the urban average, suggesting factors such as poverty further add to the heterogeneity. However, in some countries, as a result of targeted, local interventions, the opposite influence was seen where rural dwellers had easier access to healthcare providers they trusted. In many cases, adjusting for wealth and health care access and travel costs diluted the urban-rural differential, suggesting it is not the driving factor but rather a description of other characteristics. As a result of this variation, we found no significant pooled effect in our meta-analysis.

A number of contributing demographic factors were highlighted; however, their influence on coverage varied by setting and country. We found a significant relative risk to being fully vaccinated given female compared to male, suggesting females were 3% (95%CI [1%,5%]) less likely to be fully vaccinated than males. Gender disparities were identified in a number of countries and this was exacerbated by other factors, including poverty. In the two included studies with a pro-female vaccine heterogeneity, overall disparity was low or otherwise explained, suggesting that gender inequality could result from conditions of inequality in other areas. Other demographic factors such as ethnicity, caste, and religion were noted to influence vaccine coverage achieved. In India and Nepal, children in lower castes were less likely to be fully immunised, similarly, those in minority religious groups were less likely to be fully immunised in Burkina Faso, Cameroon, Ethiopia, Kenya, India and, to some extent, Nigeria [42, 98, 56, 101, 100, 57]. Lastly, birth order, family size and mothers’ marital status were all highlighted in some studies as being influential but the overall picture was mixed. In the meta-analysis we found no significant pooled effect of mother’s marital status on child immunisation. Higher birth order and larger family size had a positive effect on full immunisation coverage in China, Swaziland, and Tanzania, but a negative effect in India, Indonesia, Kenya, Mozambique, Nigeria, and Pakistan suggesting other contributory factors.

Socioeconomic factors such as household wealth, parental education, and/or occupation were some of the most significantly influential considerations for full immunisation coverage. We found a significant relative benefit of being in the highest wealth quintile for full immunisation coverage, with the wealthiest quintile 82% (95%CI [40%,137%]) more likely to be fully vaccinated than the poorest. This influence was almost unanimous across the studies; only four indicated a contrasting trend possibly motivated by outreach activities or other confounding factors. Parental occupations are often included in the wealth metric and have been found to contribute individually to the likelihood of child immunisation but only in select countries e.g. Nigeria, Pakistan, and Bangladesh. In contrast, parental education, particularly of the mother, was found to be a significant factor in child immunisation. We estimated that children were 27% (95%CI [16%,36%]) more likely to be fully vaccinated if their mother had primary level education or above, compared to no education. This trend was significant and consistent across the majority of studies although some found negligible or conflicting associations. Maternal education and access to education are often linked to economic status. One study showed that of the women in the lowest wealth quintile 64% had no formal education and a further 52% of women in the poorer wealth quintile had no formal education [102]. Vaccination coverage may be higher for children whose mothers are more educated, as these mothers may be in a better position to understand the importance of vaccination. [123]. Similarly, wealth, education, and media consumption could inform trust and knowledge regarding vaccines which may affect healthcare seeking behaviour. This can be seen in the link between access to media and an increase in vaccination rates [124]. A summary of all meta-analysis results can be found in supplementary table A.1.

We found few reviews with the scope of our research question in LMICs. A systematic review of equity in India by Mathew found similar underlying trends to those we report here [125]. The review noted a disparity in achieved coverage between urban and rural areas with 57.6% or 38.6% of infants immunised respectively and noted that boys had a higher vaccination rate than girls, by 3.8 percentage points, and that maternal literacy, often used as a proxy for education, had a positive influence on childhood immunisation. Finally, it found that the urban poor were disadvantaged in terms of vaccination achieved and that vaccine dropout was a known issue for disadvantaged populations. Similarly, a review in Nigeria by Williams, Akande, and Abbas also highlighted that living in rural areas and poorer households with no formal education or antenatal visits contributed to lower vaccination coverage achieved [126]. A review in 64 LMICs found that ethnic disparities resulting in increased zero-dose prevalence persist in the majority of countries, even when adjusting for other major sociodemographic factors; notable exclusions were Angola, Benin, Nigeria, and the Philippines [127].

When we examine similar research into equity in coverage in high income countries (HICs), we see some contrasting relationships. Arat et al. reviewed studies examining European countries and Australia and noted no significant contribution of socioeconomic factors to vaccine coverage heterogeneity in the majority of studies. This is in sharp contrast to our own results which highlight socioeconomic factors as being particularly influential. They also examined maternal education and found mixed influence on childhood immunisation, whereas we found maternal education to be significantly beneficial [128]. These results highlight the nuanced differences in inequality internationally.

The COVID-19 pandemic has placed a great strain on existing health systems globally, increased health and vaccine inequities, and prevented many from being able to access key health care services. This is in part due to non-pharmaceutical interventions (NPIs), fear of contracting COVID-19, staff absences, and the redirection of resources towards COVID-19 response services. This has greatly impacted the delivery of care services with reductions seen in vaccination coverage; for example, Brazil saw a 20% drop in coverage, particularly in poorer socioeconomic areas, and Bangladesh saw a 50.4% reduction in children immunised in April 2020 [129, 130, 131]. Delays or disruption to immunisation can lead to outbreaks and disease resurgence. This was previously observed in the Democratic Republic of the Congo (DRC) where the 2018 Ebola outbreak led to a resurgence of measles, and is projected to occur in the situation of COVID-19 disruption with respect to other outbreak-prone pathogens [129, 132].

Whilst vaccination against COVID-19 was not included in the current study, we may expect this rollout to have complex implications for existing vaccination programmes. Vaccine nationalism and scarcity have affected the deployment of COVID-19 vaccines [133]. Some HICs are currently offering booster doses whereas some LMICs, with large at-risk populations, have yet to receive initial doses [134, 135]. Vaccine nationalism has negatively influenced the supply of doses through COVAX and intellectual property protection has created a barrier to access for the majority of countries [136, 137].

One limitation of this study is that many of the studies included used secondary data which may have been collected for another primary purpose. There may additionally be publication and recall bias as the majority of studies utilised Demographic and Health Surveys (DHS). These surveys often used vaccine cards to collect data. However, where vaccine cards were unavailable, the data relied on parental (often mothers’) recall of the vaccinations received. This introduced bias to the data sets particularly data where a larger percentage of children did not have a vaccination card. There may also be a potential English language bias that impacts the data the studies used for this systematic review. Most of the studies conducted were from Asia, and East and South Africa with little to no country-specific studies from North African, Middle Eastern, European, or South American countries. A potential source of upwards bias is that secondary data from DHS depended on data from census maps, which may be outdated or incomplete. Across the countries and studies, the included vaccines and age ranges varied. Our meta-analyses we only compares fully vaccinated with not fully vaccinated individuals, leading to the removal of Uthman et al. from the quantitative results. However, in the thematic analyses we include all definitions, vaccines and age groups which may lead to vaccine-specific effects being missed. This also leads to the differing definitions of delayed vaccination. Further, the metric of wealth quintiles is a source of extensive classification and research; as such, there may be variations in definition used between studies. In the meta-analysis, we compare like for like per study; however, in the pooled results or thematic analysis, this factor may be more influential. Similarly, we compare across countries and time periods in order to understand possible themes across all LMICs; however, there is notable heterogeneity in some cases, as we have highlighted regarding urban-rural differences. Finally, despite our exclusion of COVID-19 vaccines to examine long-standing inequities in immunisation coverage, we note that the recent imbalanced rollout of COVID-19 vaccines has led to unique inequalities worthy of their own study.

## 5 Conclusions

Our findings indicate that sociodemographic determinants of health are major barriers to vaccine equality in LMICs, with vaccine inequality leading to increased morbidity and mortality. Globally, considerable progress has been made in increasing vaccine equity through global policy initiatives, including the UN’s SDGs, the GVAP, and IA2030. The stagnation of progress in DTP3 coverage since 2010, and the more recent disruption of routine immunisation services and campaign activities as a result of the COVID-19 pandemic, will result in an estimated 5% fewer vaccinated persons and 5.22% more vaccine-preventable deaths for vaccination activities occurring between 2020 to 2030, even when IA2030 goals are met [6]. Further, the recent downward trends in funding for immunisation programmes, despite the issues raised by the global COVID-19 pandemic, mean the national prioritisation of immunisation remains crucial. Reducing vaccine inequalities will thus require stronger global commitment to international immunisation targets and the implementation of catch-up campaigns to address gaps in existing immunity.

## Data Availability

Data and code are available from https://github.com/mrcide/vaccinequity_litreview.

https://github.com/mrc-ide/vaccinequity_litreview.

## List of abbreviations

Glossary: 
DTP: Diptheria, tetanus and polio (sometimes shown as DPT). 2
EPI: Expanded Programme on Immunisation. 16
HIC: High income country. 22
LMIC: Low or middle income country. 2
OPV: Oral polio vaccine. 12
SDG: Sustainable development goals. 2

## Competing interests

The authors declare that they have no competing interests.

## Funding

This work was carried out as part of the Vaccine Impact Modelling Consortium (VIMC, www.vaccineimpact.org). VIMC is jointly funded by Gavi, the Vaccine Alliance, and by the Bill Melinda Gates Foundation. The views expressed are those of the authors and not necessarily those of the Consortium or its funders. The funders were given the opportunity to review this paper prior to publication, but the final decision on the content of the publication was taken by the authors. Consortium members received funding from Gavi and BMGF via VIMC during the course of the study. This work was supported, in whole or in part, by the Bill Melinda Gates Foundation, via the Vaccine Impact Modelling Consortium (Grant Number OPP1157270 / INV-009125). AMH, SEL, JR, XL, KW, NMF, JT, KAMG also acknowledge funding from the MRC Centre for Global Infectious Disease Analysis (reference MR/R015600/1), jointly funded by the UK Medical Research Council (MRC) and the UK Foreign, Commonwealth Development Office (FCDO), under the MRC/FCDO Concordat agreement and is also part of the EDCTP2 programme supported by the European Union; and acknowledge funding by Community Jameel.

## Author’s contributions

HA made substantial contributions to the acquisition and interpretation of data and substantially revised the work. AMH made substantial contributions to the acquisition and interpretation of data and substantially drafted and revised the work. SEL made substantial contributions to revisions of the work. JR made substantial contributions to revisions of the work. XL made substantial contributions to revisions of the work. KA made substantial contributions to revisions of the work. AP made substantial contributions to revisions of the work. EV made substantial contributions to revisions of the work. KW made substantial contributions to revisions of the work. NMF made substantial contributions to the conception of the work. JT made substantial contributions to the design of the work, acquisition and interpretation of data and has substantially revised it. KAMG made substantial contributions to the design of the work, acquisition, analysis and interpretation of data and has substantially drafted and revised it. HA, AMH, SEL, JR, XL, KA, AP, EV, KW, NMF, JT, KAMG approved the submitted version of the manuscript and agree to be personally responsible for their author’s contributions and will ensure that questions related to the accuracy or integrity of any part of the work are appropriately investigated, resolved, and the resolution documented in the literature.

## Ethics approval and consent to participate

Not applicable

## Consent for publication

Not applicable

## Availability of data and materials

The data and code analysed in this manuscript are available from the following GitHub repository https://github.com/mrc-ide/vaccinequity_litreview.

## Appendix A: Supplementary Material

### A.1 Age range of study vaccinees

**Figure A.1:**
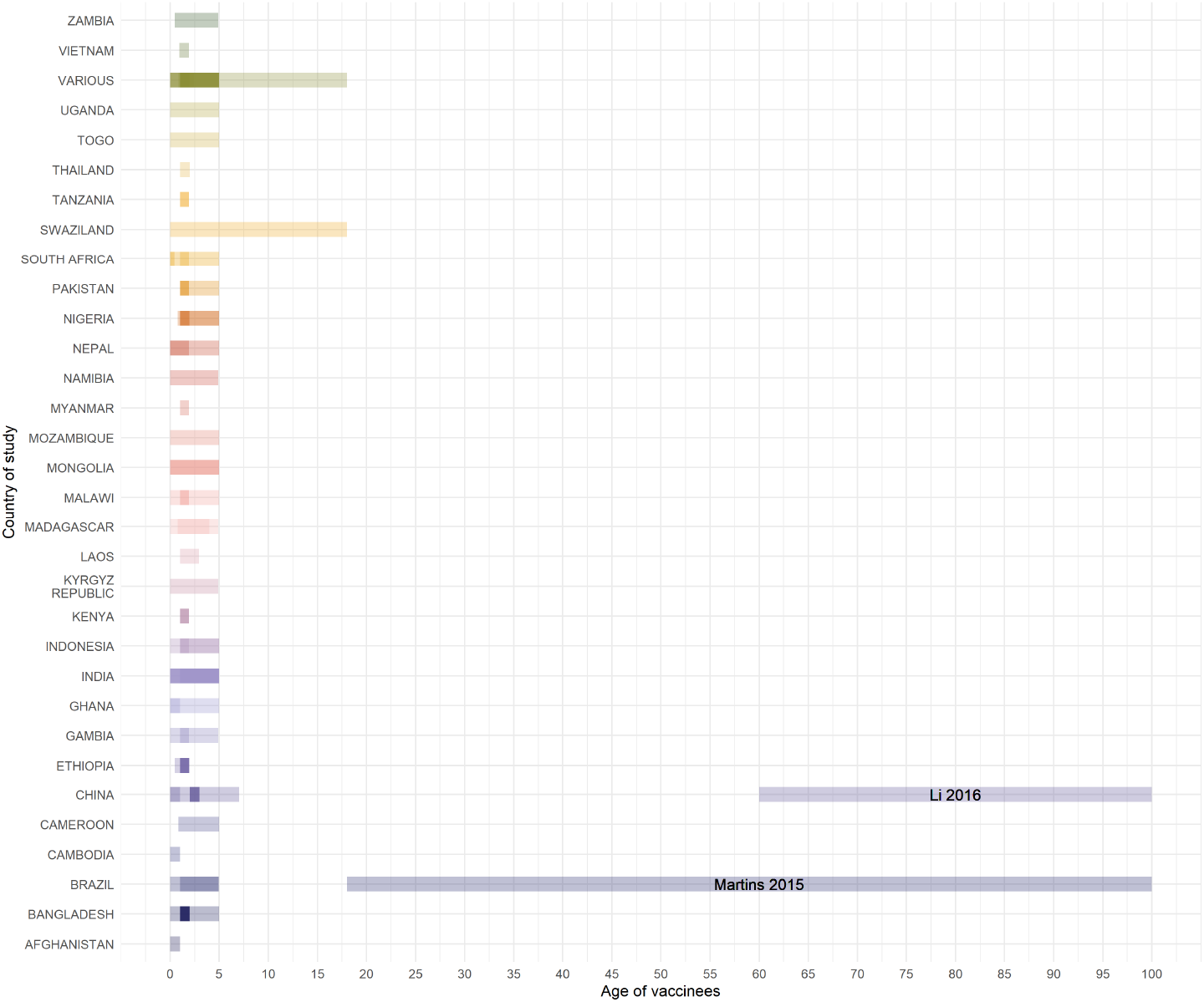
Age ranges of participants or vaccinees in included studies. The bar extends from minimum to maximum age of participants. Where multiple studies are conducted in a country, their bars overlap so darker regions indicate age groups covered by more than one study. Two studies are highlighted as they examined adults [36, 37] all other studies examined children. Three studies did not include clear age ranges and are omitted from the figure [30, 121, 41].

In the majority of cases, studies detailed the age range of the study participants who were considered for immunisation. Where this was less clear we assumed that the age range for the standard course of vaccinations was 0 - 5 years and that “children of any age” where under 18 years old. These broad assumptions only affected a small number of studies, all age ranges are displayed in figure A.1 where each bar indicates the range of a particular study, studies conducted in the same country overlap such that the darker regions indicate age groups that are examined in multiple studies.

### A.2 Additional meta-analysis results

#### A.2.1 Quantitative synthesis of maternal marital status differences in vaccination coverage

We found no significant pooled effect of maternal marital status on full vaccination coverage (A.2). Only one included study had a significant relative benefit, Muche Fenta et al. who found that a child was 8% (95%CI [6%,10%]) more likely to be vaccinated if their mother was married [24]. However, no other study confirmed this trend and Joseph et al. found the opposite effect in Kenya, with children 2% (95%CI [5%, 0%]) *less* likely to be vaccinated if their mother was married [75], see figure A.2.

**Figure A.2:**
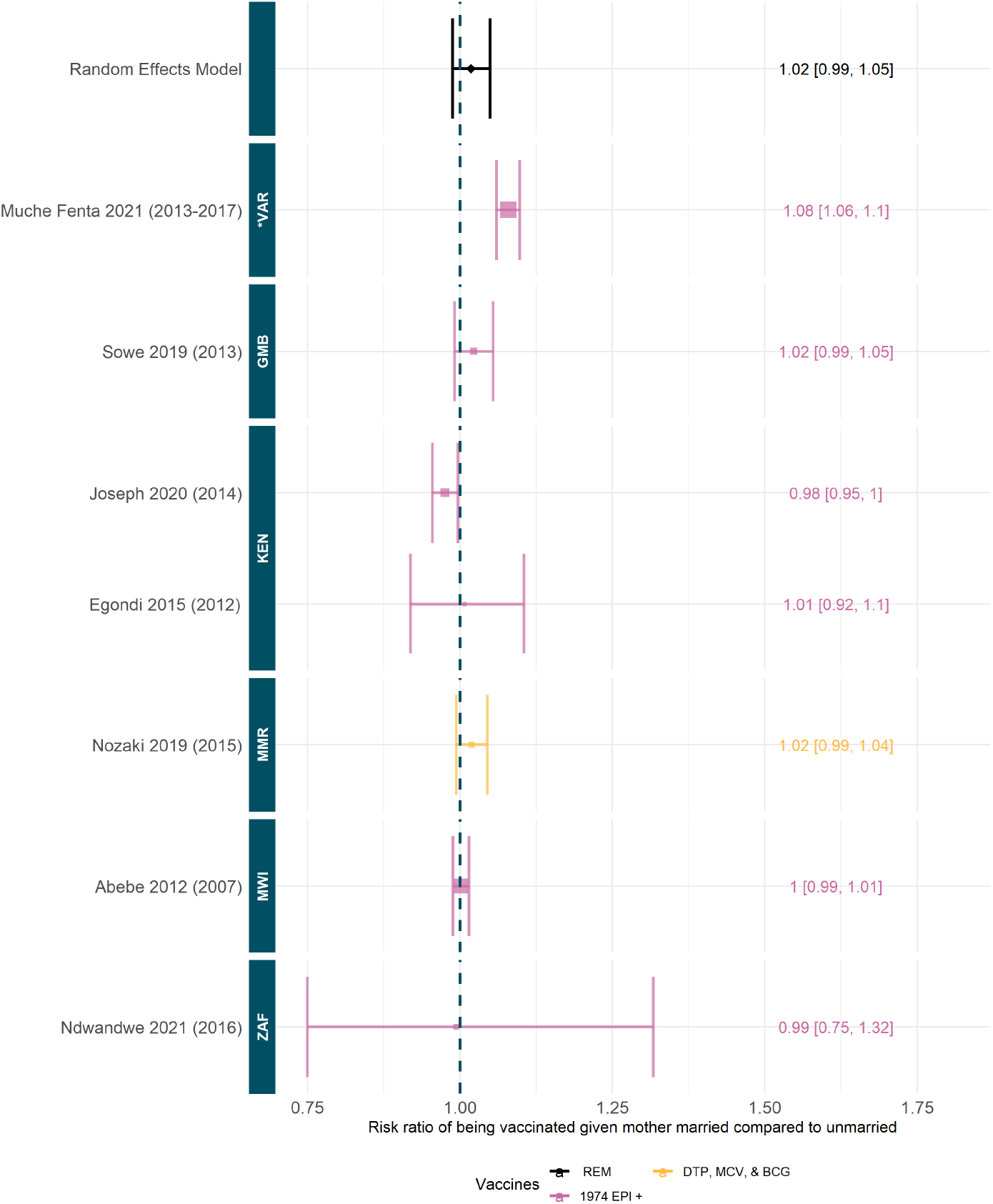
Relative risk to being fully vaccinated given mother married compared to unmarried. Random effect model estimate is shown in black and p-value of the fit is not significant (0.25). Colours denote type of vaccines considered, see table A.3 for full details. Studies are ordered by the year of data, shown in brackets, and country of data. ISO codes are: ZAF=South Africa, MWI=Malawi, MMR=Myanmar, KEN=Kenya, GMB=Gambia and *VAR=Various. Studies included: [24, 88, 94, 68, 75, 74, 50].

### A.2.2 Summary of meta-analysis Results & insights into Uthman et l. exclusion

**Table A.1:** Summary table of all meta-analysis results.

| Attribute | # Studies | Meta-analysis Result | Heterogeneity |
| --- | --- | --- | --- |
| Urban/Rural | 24 | No significant pooled effect | Wide heterogeneity between countries with, for example, all studies in China suggesting a positive risk ratio and all studies in Ethiopia suggesting a negative risk ratio. |
| Gender | 26 | Significant relative risk to being fully vaccinated given female compared to male of 0.97 (95%CI [0.95, 0.99]) suggesting females are 3% (95%CI[1%,5%]) less likely to be vaccinated than their male counterparts | General consensus between countries and time periods with almost all studies either spanning one or with significantly negative risk ratio. |
| Education | 16 | Relative risk to a child being vaccinated given their mother has no formal education compared to having any education level ie. primary or above, as 0.73 (95%CI [0.64, 0.84]); this model fit is significant. This implies that children are 27% (95%CI [16%,36%]) less likely to be fully vaccinated if their mother has no formal education. | General consensus between countries and time periods with almost all studies either spanning one or with significantly negative risk ratio. Only two studies had a significantly positive risk ratio, in the Gambia and Malawi [50, 68]. |
| Wealth | 18 | Relative risk of 0.73 (95%CI [0.63, 0.84]) to being vaccinated if in the poorest wealth quintile compared to richest; individuals in the poorest subpopulations are 27% (95%CI [16%,37%]) less likely to be fully vaccinated than those in the richest. | General consensus between countries and time periods with almost all studies either spanning one or with significantly negative risk ratio. Only two studies had a significantly positive risk ratio, in the Gambia and China [48, 50]. |
| Marital Status | 7 | No significant pooled effect | General consensus between countries and time periods, almost all studies had risk ratios spanning one. One study had a significantly positive risk ratio [24] which covered Sub-Saharan Africa. |

**Table A.2:** Relative risk estimates including Uthman et al. for low immunisation coverage [38].

| Characteristic | Relative risk | p-value | Significant given Bonferroni correction |
| --- | --- | --- | --- |
| Gender | 0.96 (95%CI[0.95, 0.98]) | $4 \times 10^{-4}$ | Yes |
| Wealth | 1.85 (95%CI[1.44, 2.38]) | $1.96 \times 10^{-6}$ | Yes |
| Urban/Rural | 1.00 (95%CI[0.93, 1.08]) | 0.91 | No |
| Maternal education | 0.72 (95%CI[0.63, 0.82]) | $1.22 \times 10^{-6}$ | Yes |
| Maternal marital status | 1.02 (95%CI[0.99, 1.05]) | 0.25 | No |

### A.2.3 Multiple Testing Considerations

Prior to data extraction, we specified 8 two-sided hypothesis tests, each of which had a null hypothesis of the form *p*_1_ = *p*_2_, where *p*_*j*_ is the probability of being fully vaccinated for subgroup *j*. We defined the two subgroups in each test (e.g. the highest quintile and the lowest quintile in the test for wealth) prior to analysing any of the data.

After data extraction, we found that we lacked the data needed to perform 3 of these hypothesis tests ^[4]^, which left us with 5 pre-specified hypothesis tests performed over the course of the analysis.

To account for multiple comparisons, our analysis uses the Bonferroni-corrected significance threshold of 0.05*/*5 = 0.01 instead of the usual value of 0.05. Using this threshold, we estimate that the probability of incorrectly rejecting the null hypothesis in at least one of our 5 comparisons (i.e. reporting at least one false positive) is 1 − (1 − .05*/*5)^5^ ≈ 0.05.

#### A.2.4 Summary of definitions of fully immunised utilised across studies

**Table A.3:** Summary table of fully vaccinated definitions across studies.

| Immunisations Included | N (%) | Studies |
| --- | --- | --- |
| 1974 EPI Vaccines (DTP, MCV, BCG, & OPV/IPV) | 41 (37.96%) | [68, 101, 54, 19, 80, 82, 69, 24, 3, 108, 88, 65, 105, 87, 62, 28, 51, 77, 116, 104, 34, 21, 22, 113, 50, 93, 86, 100, 71, 98, 109, 58, 59, 99, 64, 7, 115, 35, 42, 110, 111] |
| 1974 EPI Vaccines & HBV | 10 (9.26%) | [79, 85, 57, 107, 47, 23, 117, 60, 63, 120] |
| 1974 EPI vaccines, HBV, & MMR | 2 (1.85%) | [46, 119] |
| 1974 EPI vaccines, HBV, & JE | 2 (1.85%) | [97, 49] |
| 1974 EPI Vaccines, HBV, & Hib | 7 (6.48%) | [84, 70, 74, 72, 66, 118, 91] |
| 1974 EPI Vaccines, HBV, Hib, & YF | 2 (1.85%) | [81, 43] |
| 1974 EPI Vaccines, HBV, Hib, & MMR | 2 (1.85%) | [96, 95] |
| DTP, MCV, & BCG | 4 (3.70%) | [20, 94, 112, 61] |
| DTP & MCV | 4 (3.70%) | [8, 122, 40, 41] |
| DTP | 4 (3.70%) | [12, 26, 39, 121] |
| MCV | 11 (10.12%) | [78, 25, 55, 32, 33, 76, 67, 92, 29, 11, 30] |
| Polio (OPV/IPV) | 3 (2.78%) | [38, 27, 83] |
| HBV | 3 (2.78%) | [45, 37, 106] |
| Influenza | 2 (1.85%) | [48, 36] |
| Other | 7 (6.48%) | [75, 53, 56, 52, 90, 31, 73] |
| Not given | 4 (3.70%) | [89, 103, 114, 102] |

## Appendix B: All studies

**Table B.4:**
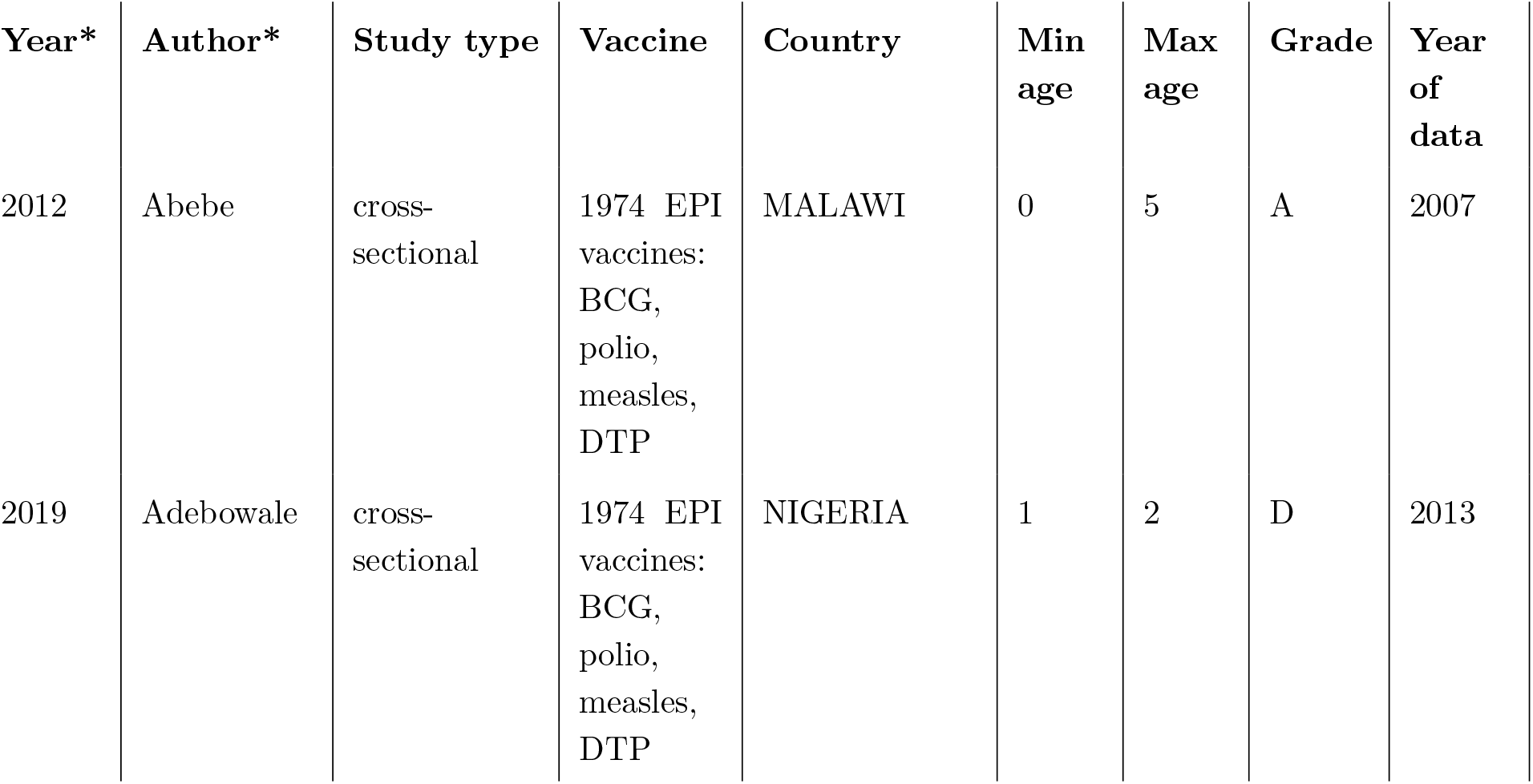
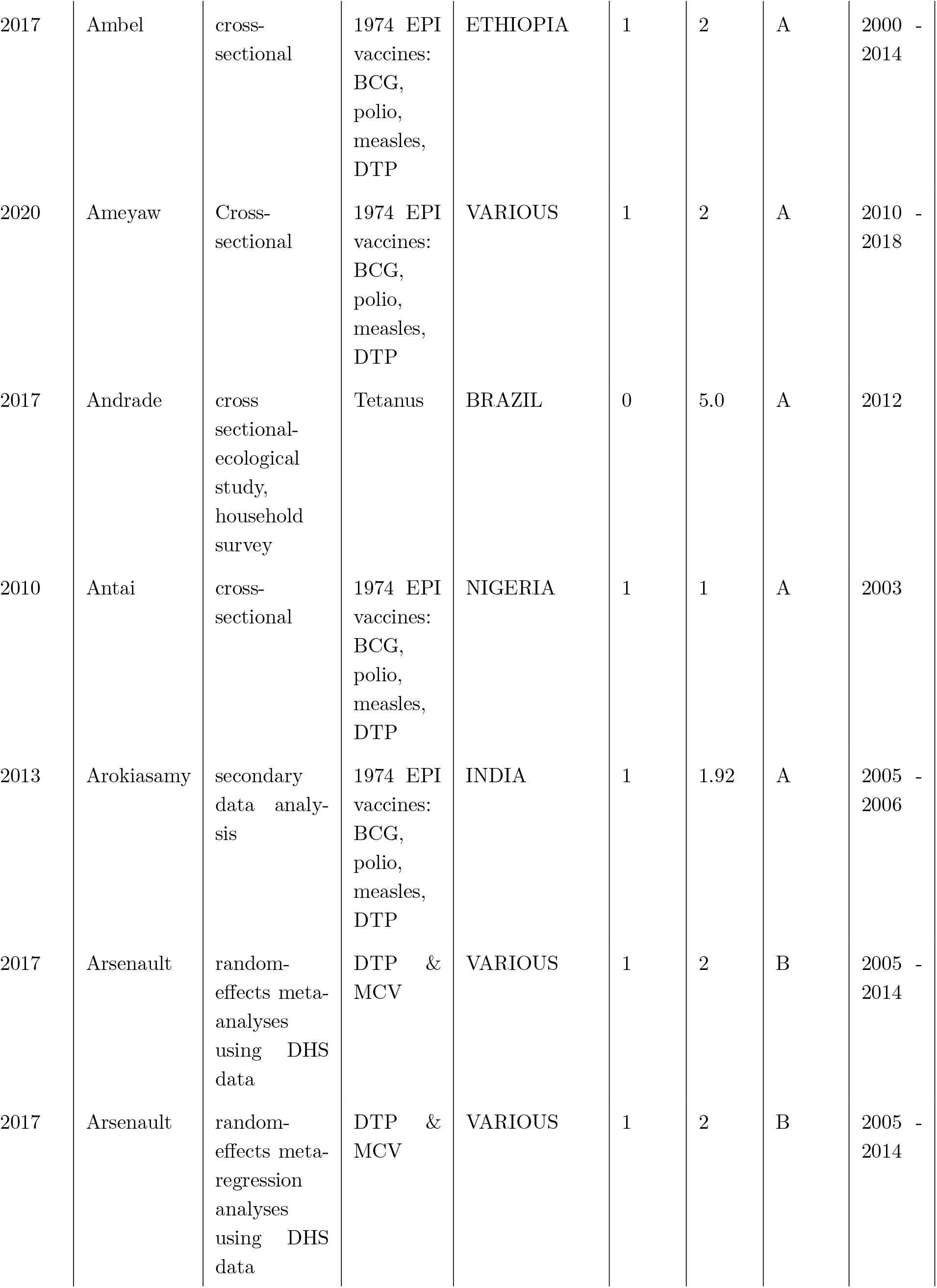
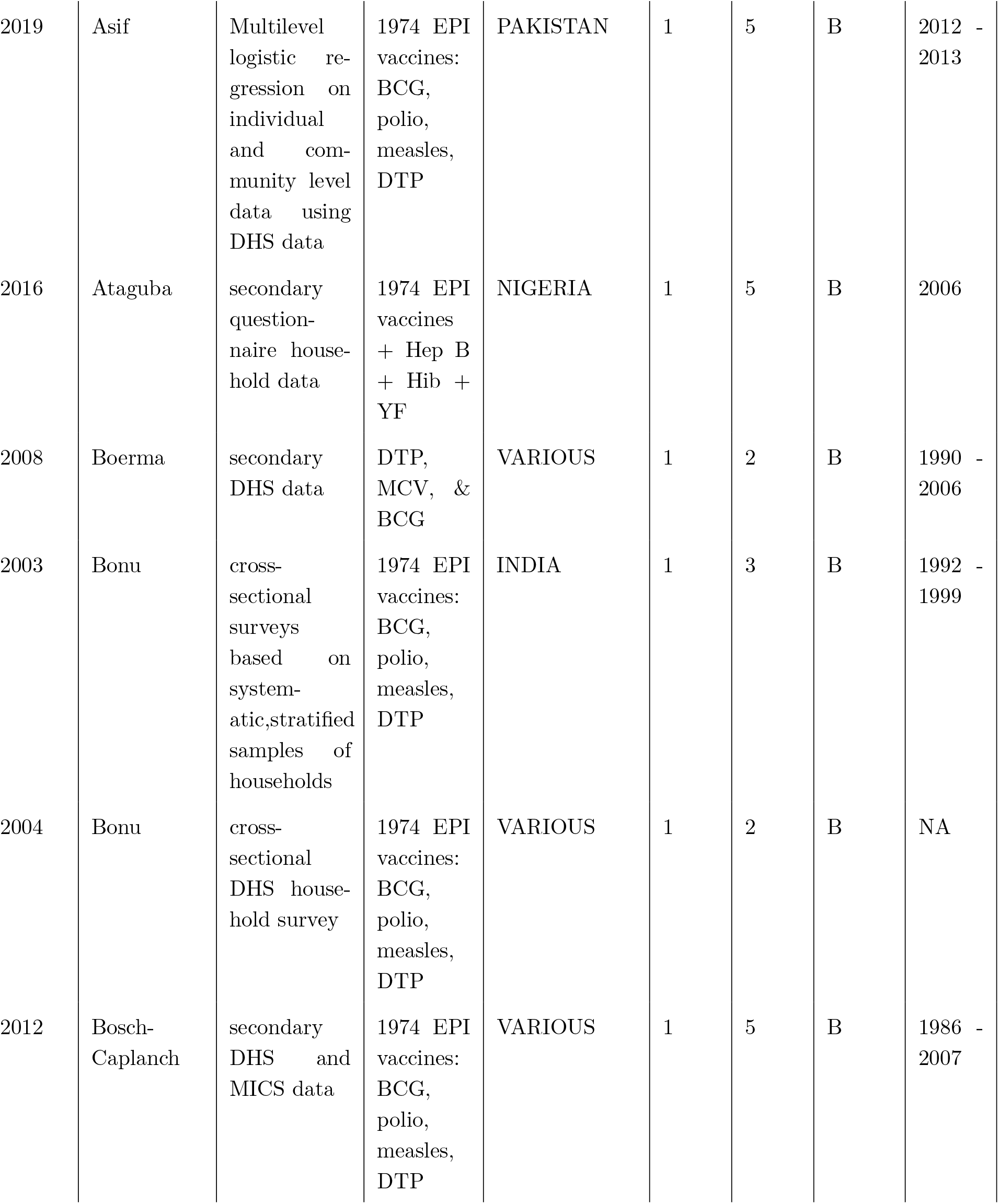
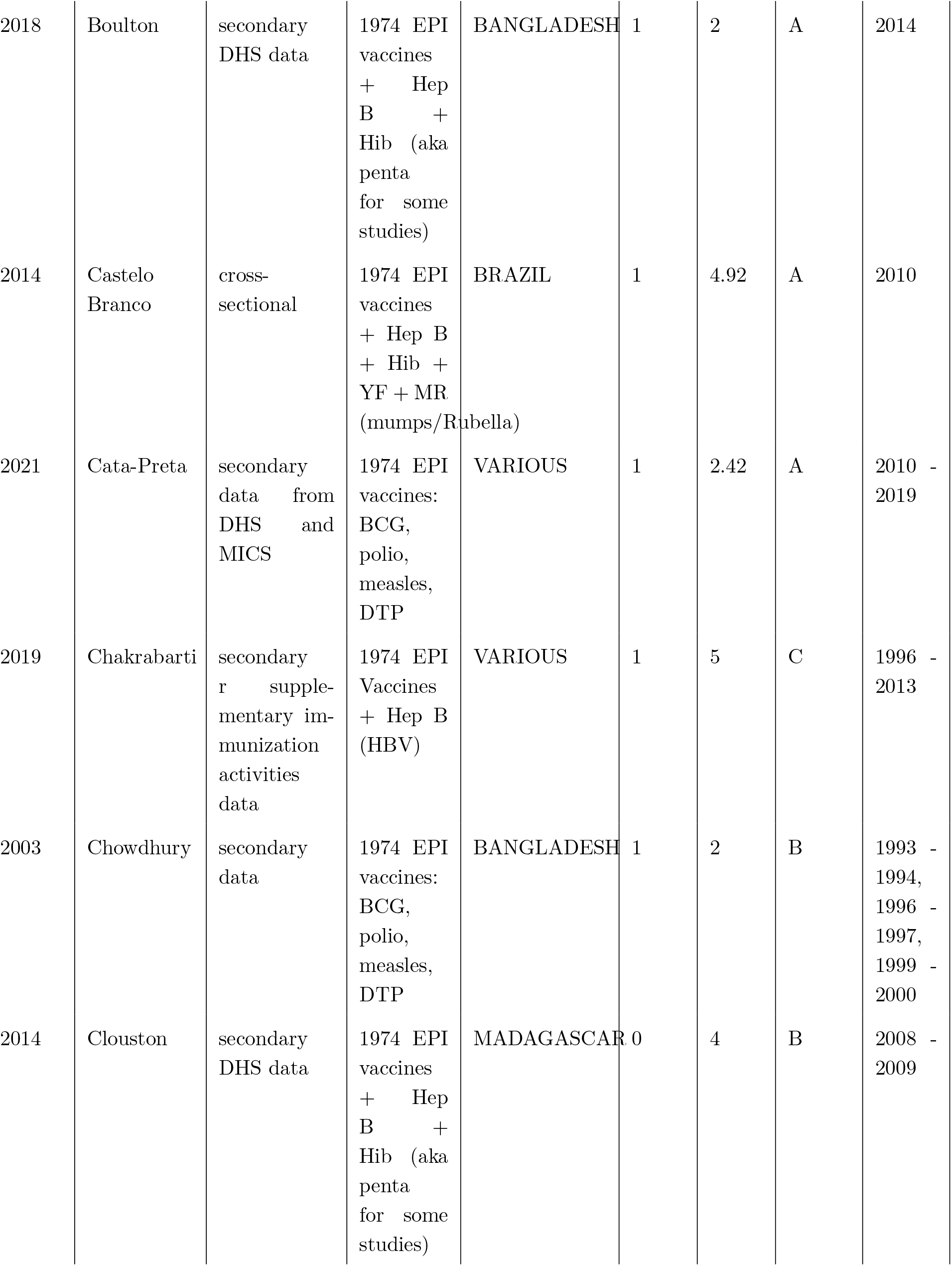
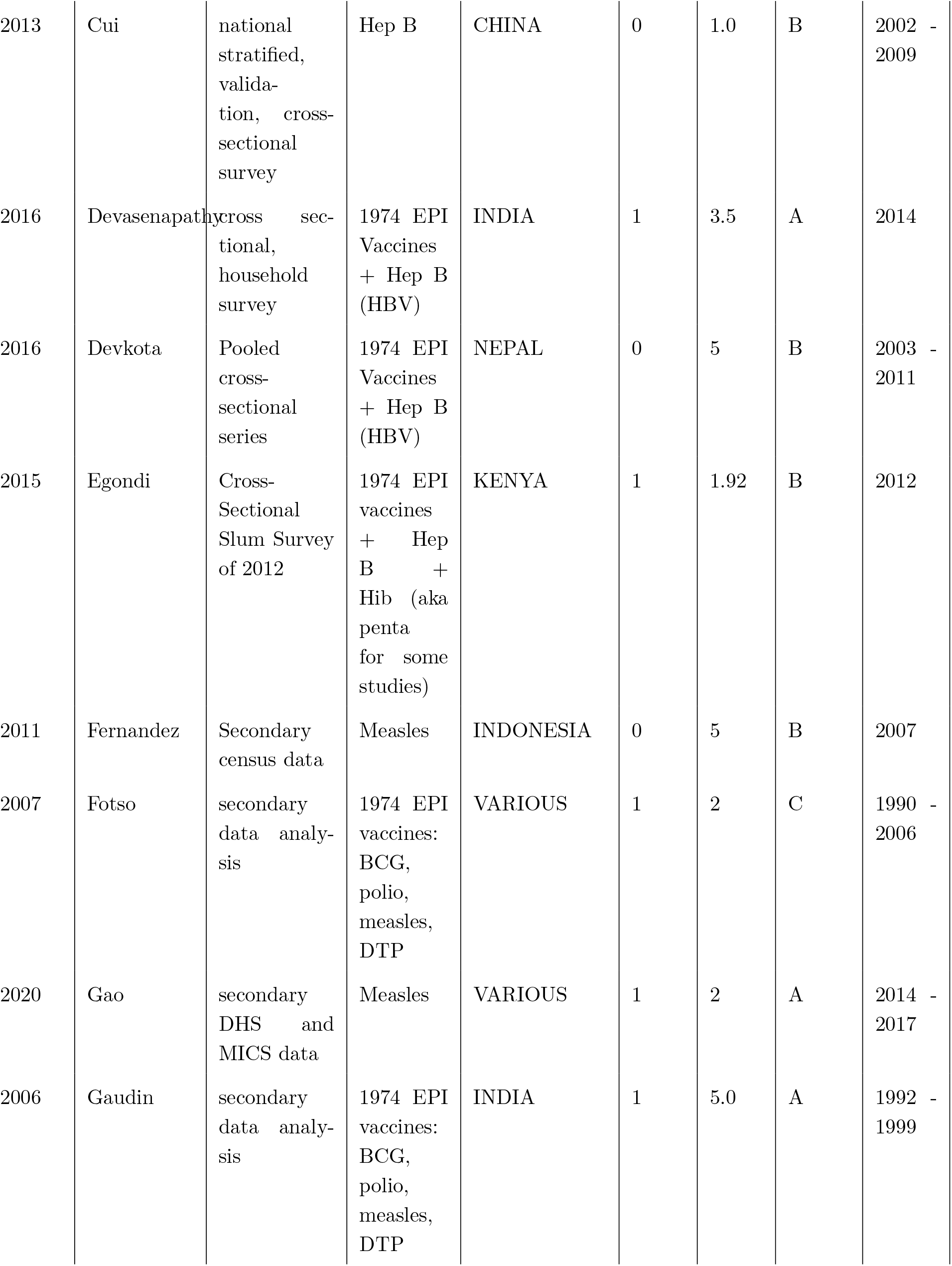
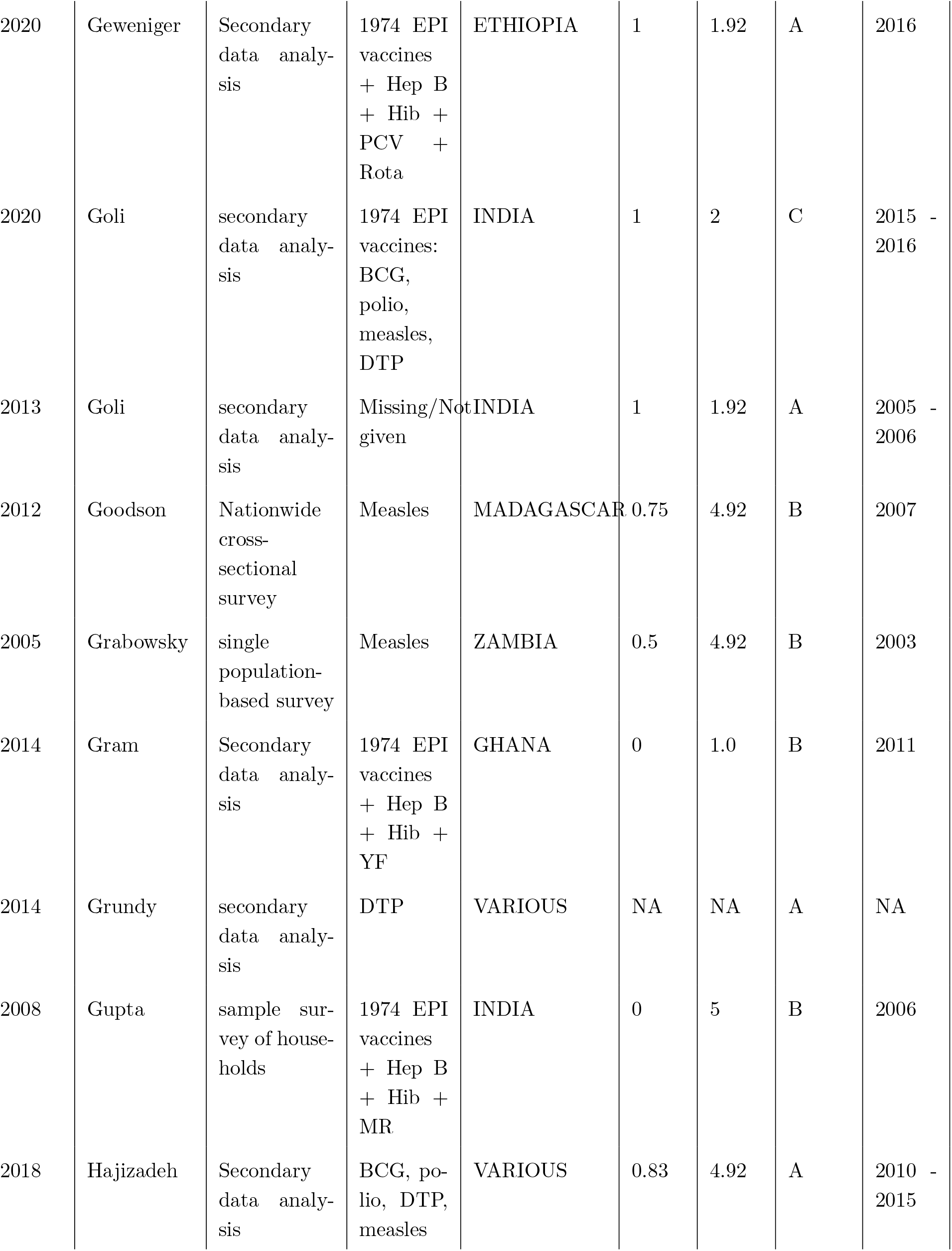
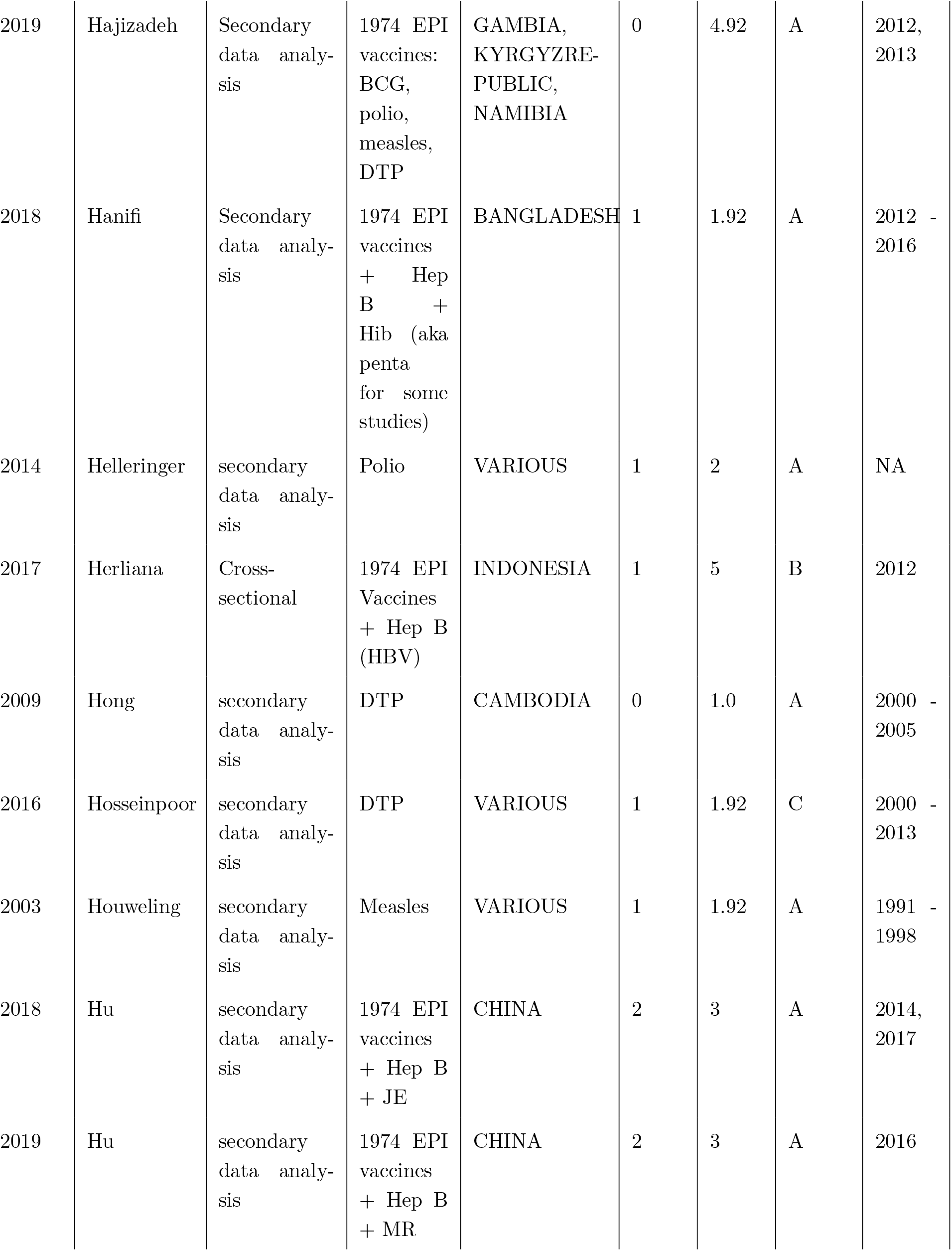
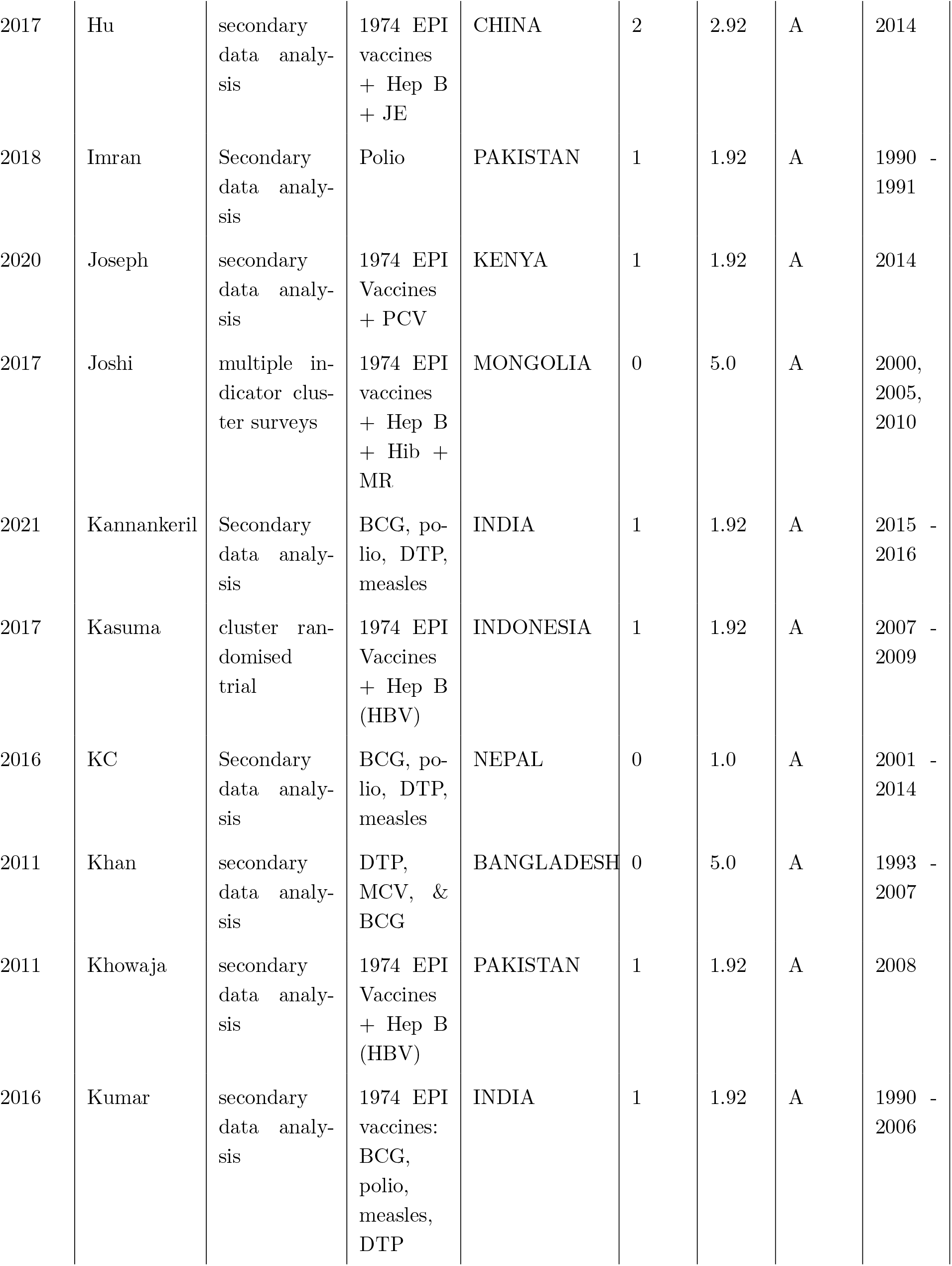
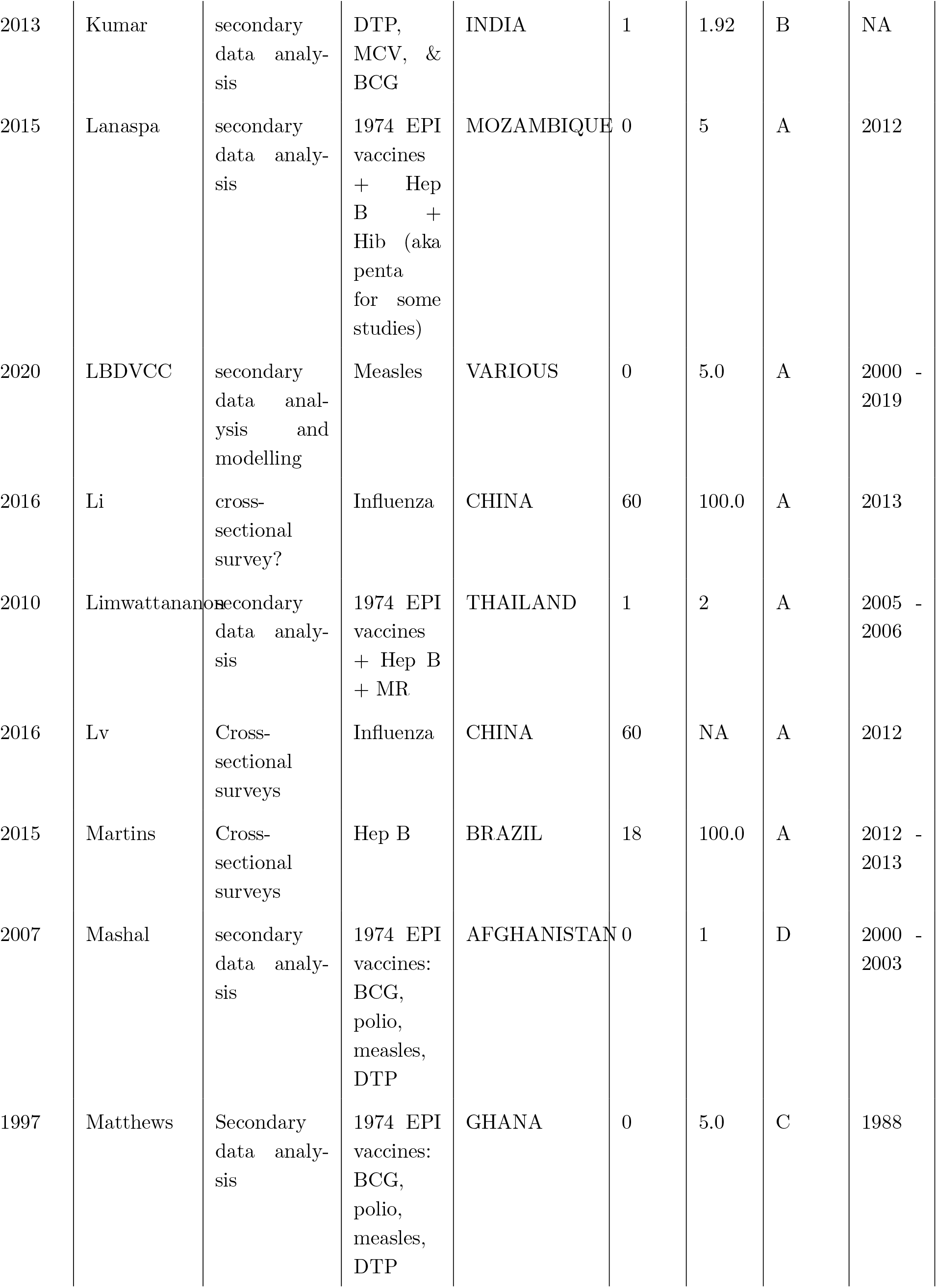
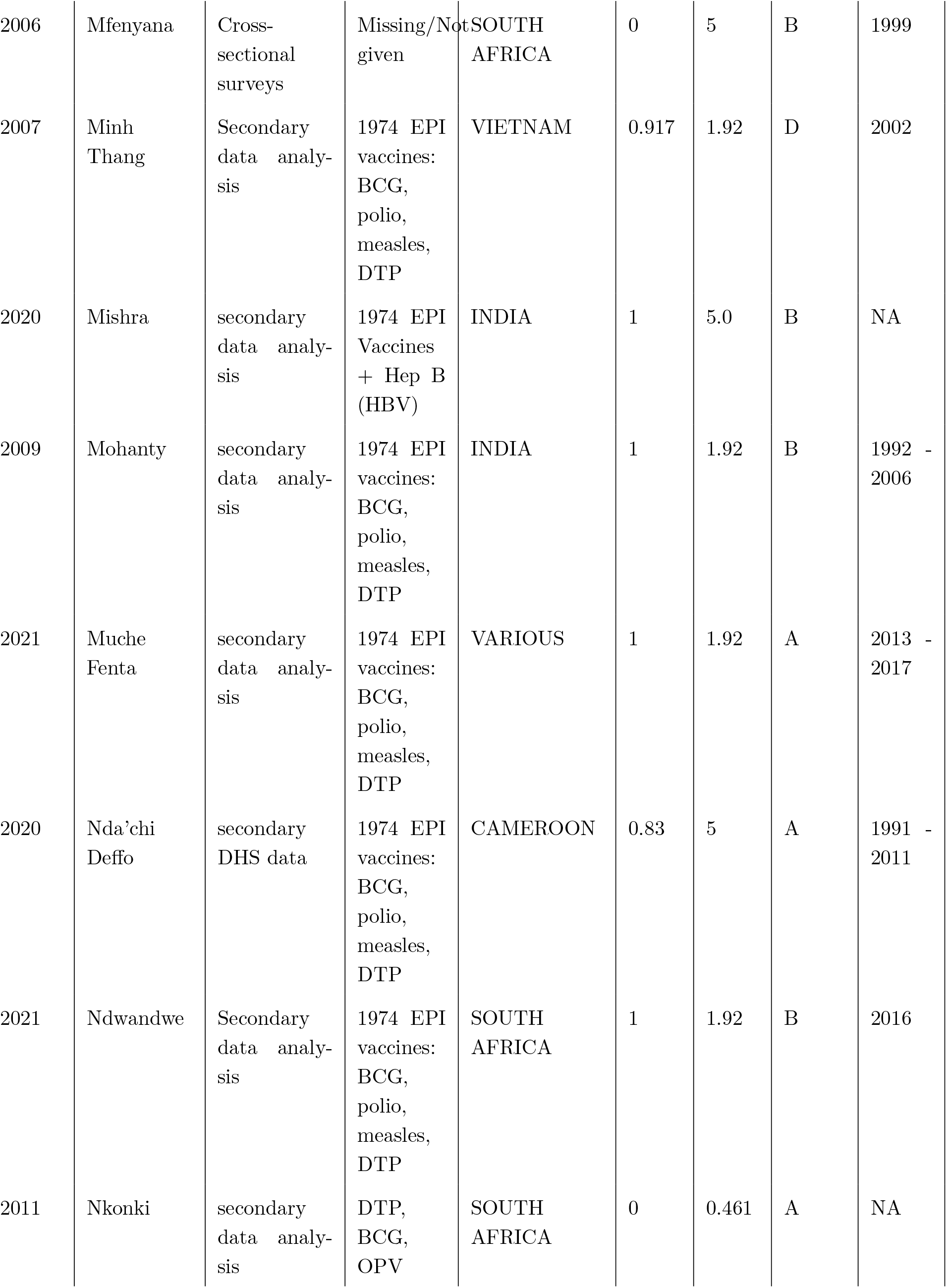
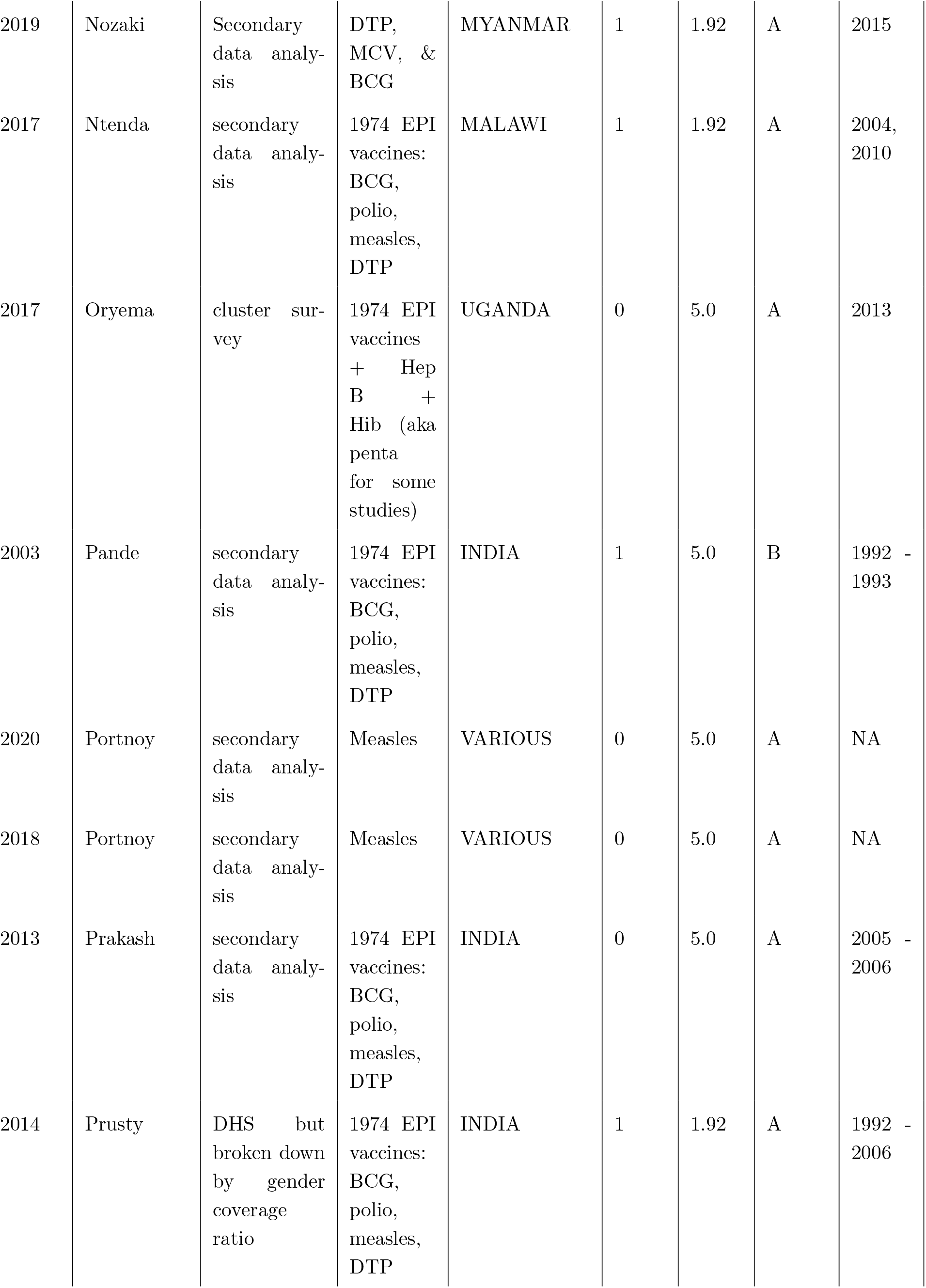
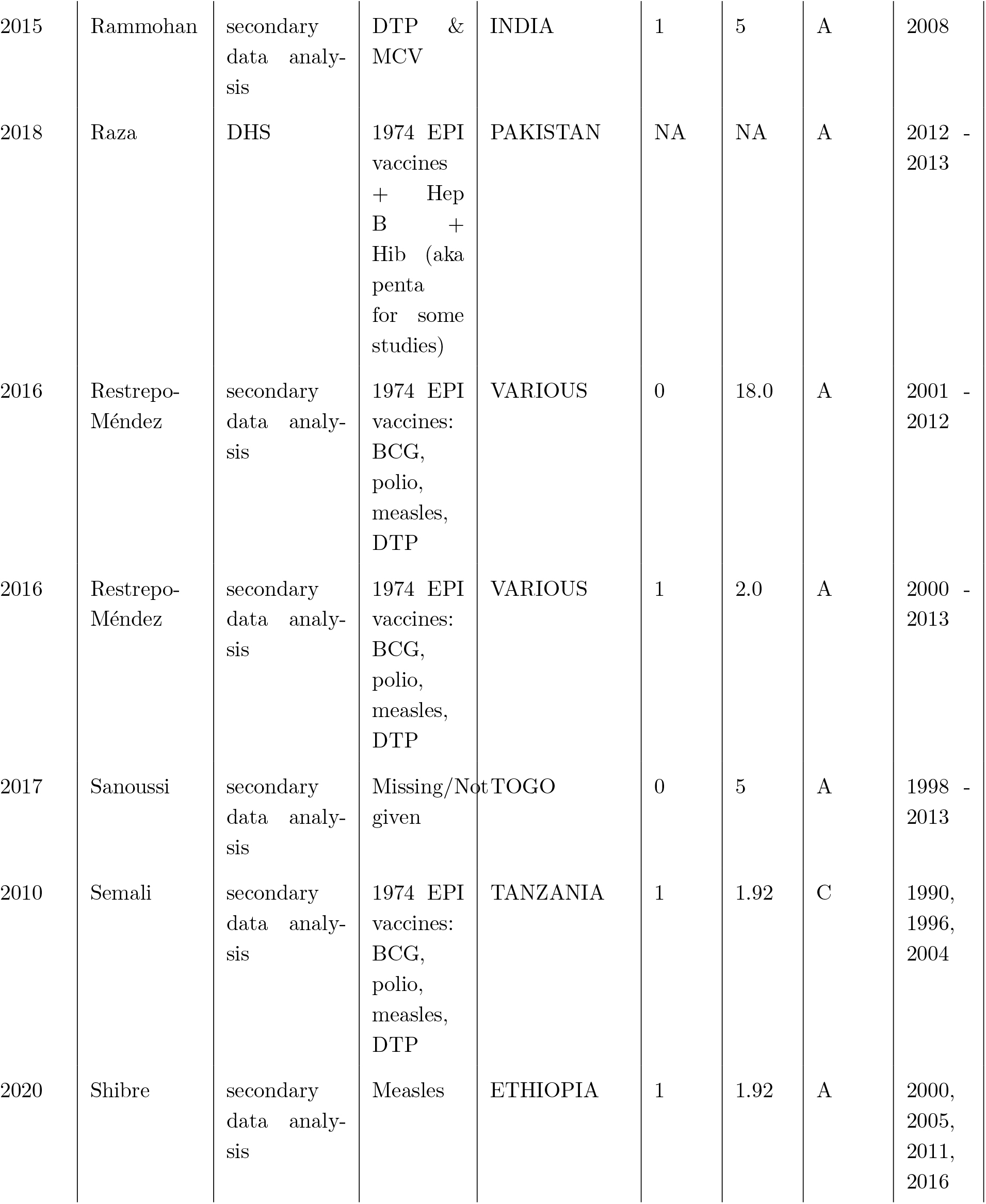
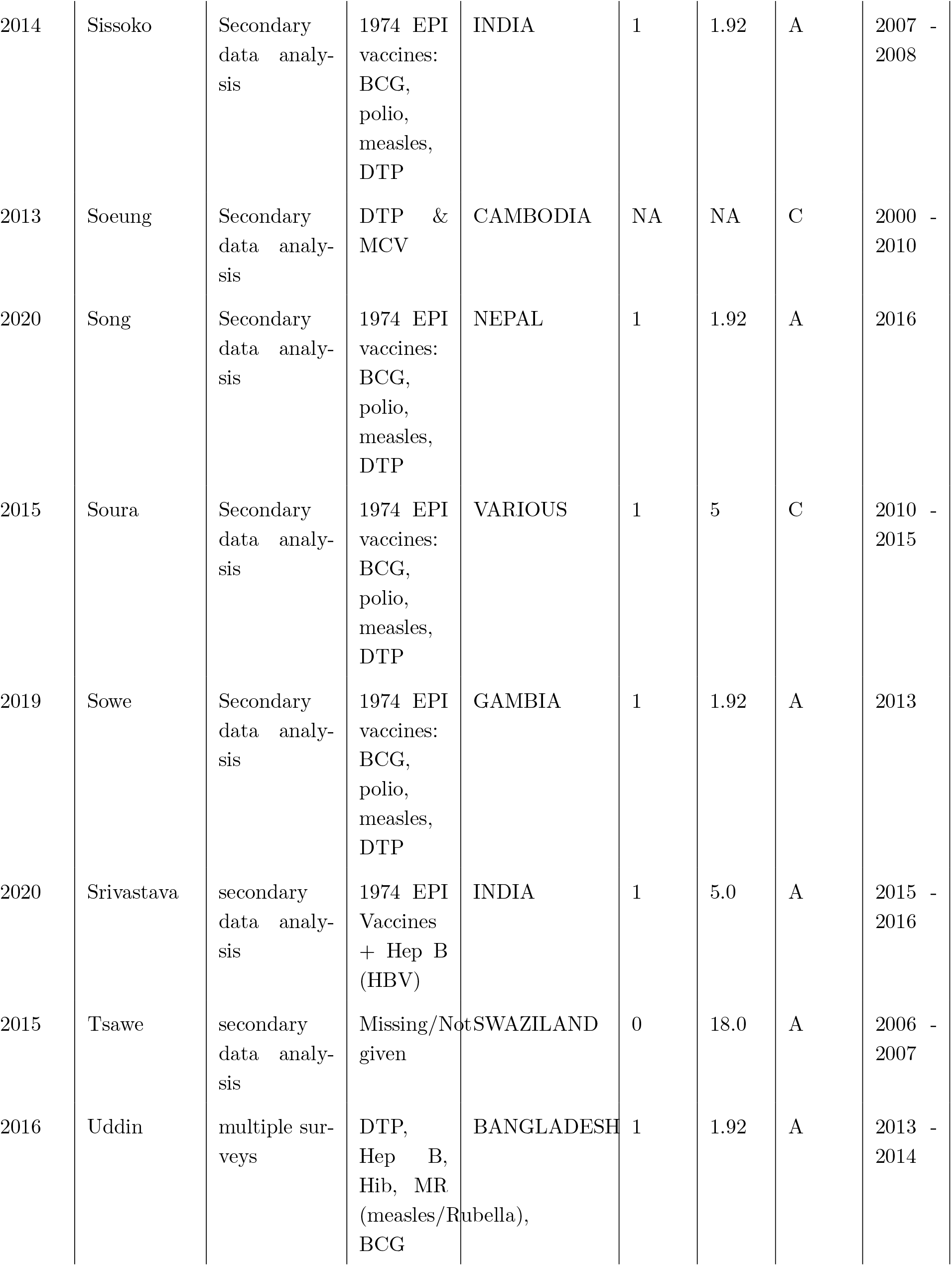
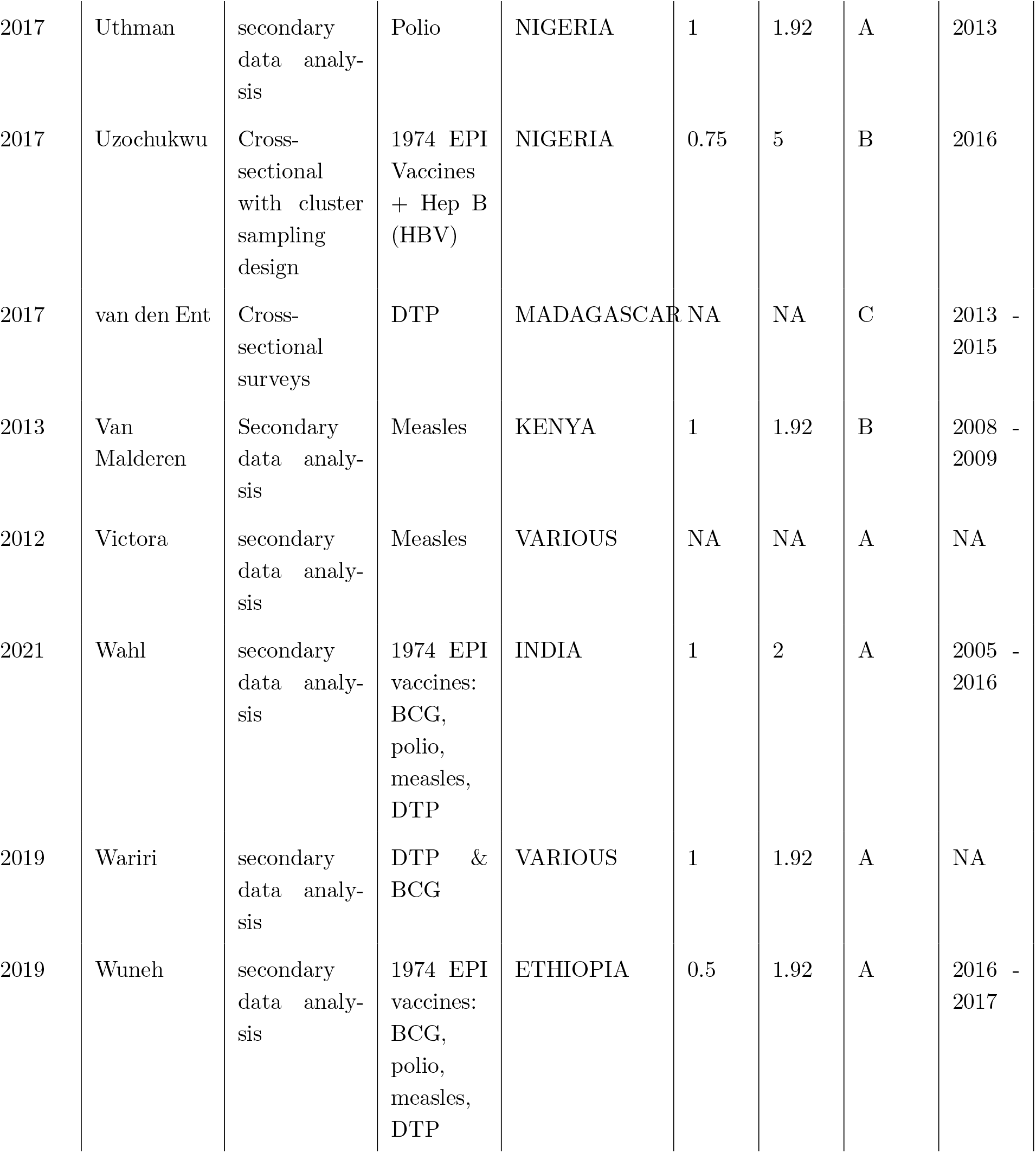
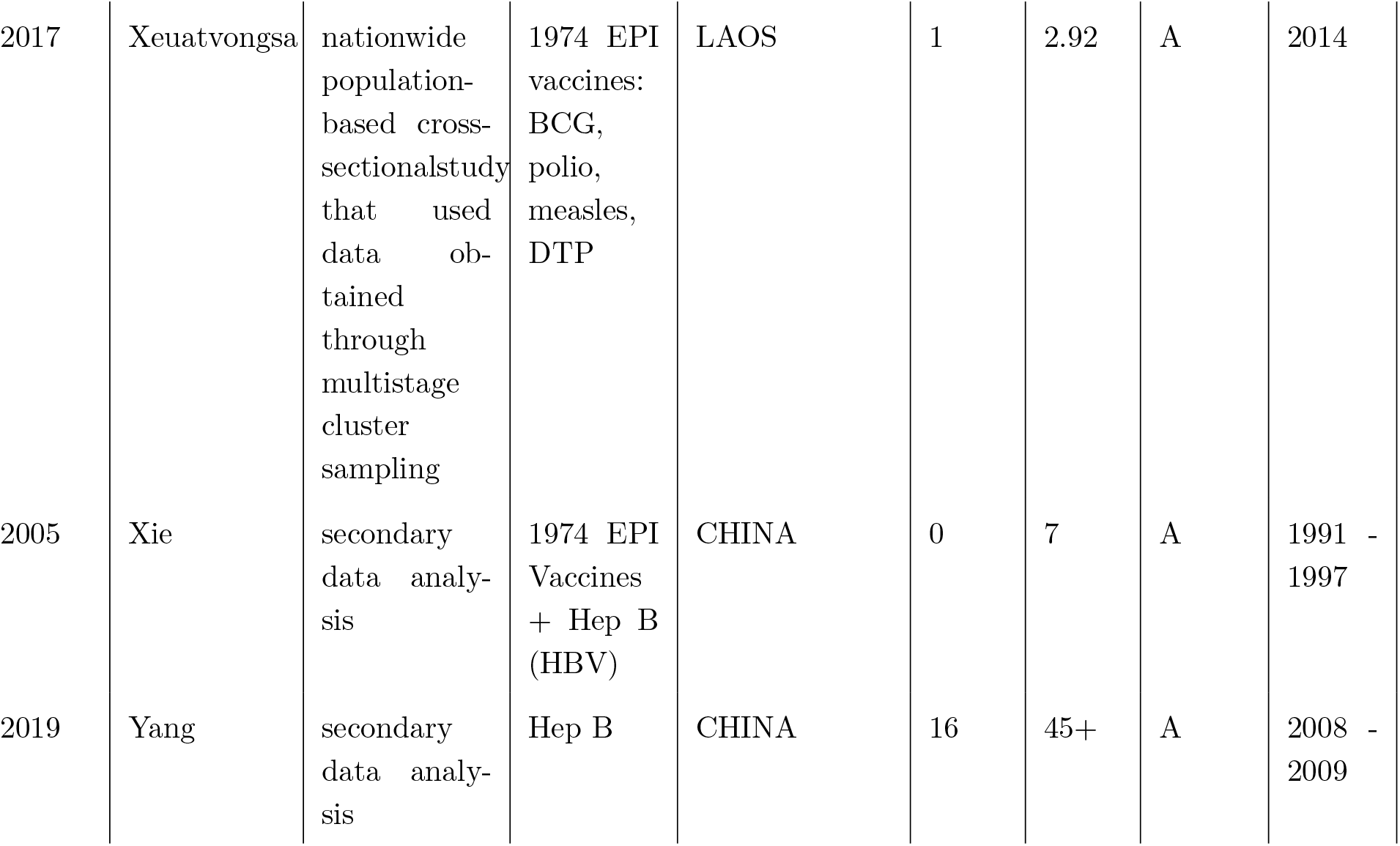
All studies used in literature review with summary characteristics. Please see the GitHub repository for all data. Year* denotes year of publication, Author* denotes first author surname.

## Footnotes

[1] Afghanistan, Angola, Democratic Republic of the Congo (DRC), Ethiopia, India, Indonesia, Iraq, Nigeria, Pakistan, and South Africa [2]

[2] ”WHO identified six major components on which the development of the immunization maturity grid was based. These components are: programme management and financing, immunization service delivery and new vaccine introduction, disease surveillance and VPD outbreak management, data management and analytics, vaccine quality, safety and regulation, and community engagement [9]”.

[3] Test were performed for the following factors: geography (urban vs rural), gender, wealth, education, and martial status.

[4] Data insufficient for variables on family size, birth order, and skilled birth attendant

## Notes

### Competing Interest Statement

The authors have declared no competing interest.

